# Epidemiology and outcomes of head trauma in rural and urban populations: A systematic review and meta-analysis

**DOI:** 10.1101/2023.10.22.23297363

**Authors:** Julia Chequer de Souza, Geoffrey P Dobson, Celine J Lee, Hayley L Letson

**Author notes:** **Email Addresses:**.

## Abstract

**Objective:** To identify and describe differences in demographics, injury characteristics, and outcomes between rural and urban head injury patients.

**Data Sources:** CINAHL, Emcare, MEDLINE, and Scopus.

**Review Methods:** A systematic review and meta-analysis of studies comparing epidemiology and outcomes of rural and urban head trauma was conducted in accordance with PRISMA and MOOSE guidelines.

**Results:** 36 studies with ∼2.5-million patients were included. Incidence of head injury was higher in males, regardless of location. Rates of transport-related head injuries, particularly involving motorized vehicles other than cars, were significantly higher in rural populations (OR:3.63, 95% CI[1.58,8.35], p=0.002), whereas urban residents had more fall-induced head trauma (OR:0.73, 95% CI[0.66,0.81], p<0.00001). Rural patients were 28% more likely to suffer severe injury, indicated by Glasgow Coma Scale (GCS)≤8 (OR:1.28, 95% CI[1.04,1.58], p=0.02). There was no difference in mortality (OR:1.09, 95% CI[0.73,1.61], p=0.067), however, urban patients were twice as likely to be discharged with a good outcome (OR:0.52, 95% CI[0.41,0.67], p<0.00001).

**Conclusions:** Rurality is associated with greater severity and poorer outcomes of traumatic head injury. Transport accidents disproportionally affect those travelling on rural roads. Future research recommendations include addition of prehospital data, adequate follow-up, standardized measures, and sub-group analyses of high-risk groups, e.g., Indigenous populations.

## Introduction

Head trauma is a leading cause of morbidity and mortality worldwide and is associated with significant healthcare costs (1, 2, 3, 4). The etiology of head trauma is varied, with vehicle accidents, falls and assaults the most common causes. Patients with traumatic head injury are at increased risk of both short- and long-term mortality and morbidity including cognitive and psychiatric disturbances, reduced quality of life, and permanent disability (5, 6, 7, 8). Several high-risk populations have been identified, including young males and Indigenous people (5, 9, 10, 11, 12).

Another important risk factor is rurality. Within Australia, one-third of the population live in rural areas, and head injuries are the most common injury requiring medical transfer (13). Similarly, an increased incidence of head trauma has been reported in rural populations in North America (14), Asia (15), Europe (16), and Africa (17). What is less clear, however, is differences in outcomes after head trauma for rural and urban patients. While some studies have reported increased mortality in rural areas compared to urban areas (18, 19), others have reported no difference (16, 20), or reduced mortality (21, 22).

Understanding the burden of head trauma and key rural/urban differences is essential to improve patient outcomes, for example, through targeted prevention strategies. The aim of this systematic review and meta-analysis is to identify and describe the differences in demographic and injury characteristics, clinical features, and outcome trends between rural and urban traumatic head injury patients worldwide.

## Materials and Methods

### Systematic review

This systematic review was conducted and is reported using the Preferred Reporting Items for Systematic Reviews and Meta-Analyses (PRISMA) guidelines (Appendix 1) (23). The protocol was registered and published with PROSPERO, an international register for systematic reviews (CRD42022336874).

### Search strategy

All studies that reported epidemiology and outcomes of traumatic head injuries with rural and urban comparisons were included. An independent literature search was conducted in CINAHL, Emcare, MEDLINE and Scopus for publications available up to February 7, 2022. The search strategy is outlined in Appendix 2. Reference lists of studies that were retrieved in full text were hand-searched to identify additional studies, and where necessary, authors of identified studies were contacted for access to full-text articles or additional data. There were no limitations placed on study size or data of publication. Studies published in a language other than English, review articles or commentaries, case studies, conference abstracts, Letters to the Editor, and animal studies were excluded. Studies were not eligible if only one mechanism of injury was analyzed, if no epidemiological data other than outcome was reported, if there was no rural and urban comparison, or if other traumatic injuries, such as cranio-facial or spinal injuries, were not reported separately from head injuries.

### Study selection

Following removal of duplicate studies, two investigators independently performed title and abstract screening to identify eligible articles. Full texts of eligible studies were retrieved and reviewed by the investigators, and a third investigator was consulted in the case of disagreement.

### Data extraction

Data were extracted for general characteristics (authors, year, title, journal, publication type), study characteristics (design, follow-up, sample size, patient source, location, definition of rurality, eligible patient identification), patient characteristics (age, sex), injury characteristics (mechanism of injury, injury severity, confirmed pathology, clinical symptoms) and outcome data (mortality, length of hospital stay, discharge status).

### Quality assessment

A modified Newcastle-Ottawa tool for quantitative research was used for quality assessment (Appendix 3). The tool included assessments for the following characteristics: representativeness of the study cohort, reporting of demographic data for both rural and urban populations, reporting of incidence/prevalence as well as outcome data, statistical methodology, inclusions of method used for determining injury severity and rural classification, and duration of follow-up. Each study was assessed as low, moderate, or high quality.

### Meta-analysis

The meta-analysis was conducted in accordance with the Meta-Analysis of Observational Studies in Epidemiology (MOOSE) guidelines (Appendix 4) (24), using Review Manager software (V5.4.1) and a random-effects model. Medians and interquartile ranges (IQR) were converted to means and standard deviations (SD) using the methods and calculator of Wan *et al* (25). Statistical significance was defined as p<0.05. Heterogeneity was determined by a significant Chi^2^ and the I^2^ statistic (I^2^ <25% low, I^2^ = 25-50% moderate, and I^2^ >50% substantial heterogeneity).

### Ethics approval

No ethical approval is required because data retrieved and analyzed was from previously published studies in which informed consent or a waiver of consent was obtained by the primary investigators.

## Results

### Study characteristics and quality assessment

A total of 1,310 studies were evaluated for rural/urban differences in head trauma patients (Fig 1). After title and abstract screening, 90 full-text articles were reviewed. No additional articles were obtained through reference list searching. Six articles were unable to be retrieved despite requests submitted to the authors. Based on our eligibility criteria, a total of 36 studies were included in the meta-analysis, representing 35 different study populations across 14 countries (Fig 1, Table 1). Most of the studies were population-based retrospective cohort studies of ≥1 year in duration. Of the 36 studies, 15 were conducted in North America (14, 18, 20, 26, 27, 28, 29, 30, 31, 32, 33, 34, 35, 36, 37) and nine in Australia and New Zealand (5, 6, 21, 22, 38, 39, 40, 41, 42). Additional information on study data sources, inclusion/exclusion criteria, and rural/urban classifications, can be found in Appendices 5-7. As per the quality assessment, 17 studies were identified as high quality (6, 14, 16, 19, 20, 29, 30, 32, 34, 37, 38, 39, 40, 41, 42, 43, 44), with the remainder determined to be of moderate quality.

**Figure 1:**
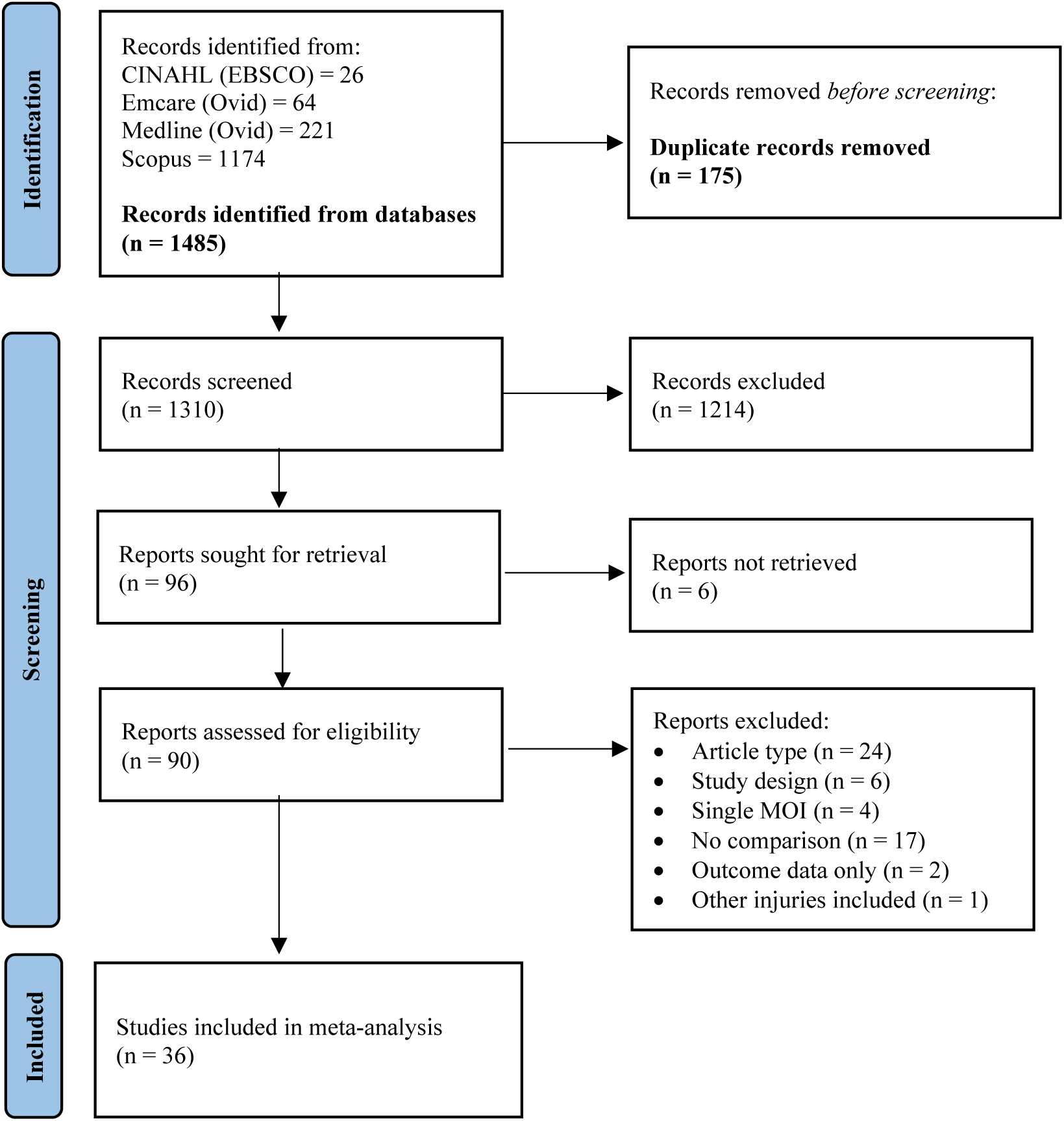
PRISMA Flow Diagram of the study selection process. A total of 1,310 studies were evaluated for rural/urban differences in head trauma patients. After title and abstract screening, 90 full-text articles were reviewed, 36 of which were included in the systematic review and meta-analysis after exclusions. CINAHL, Cumulative Index of Nursing and Allied Health; EBSCO, Elton B. Stephens Company; MOI, mechanism of injury.

**TABLE 1:** Study Characteristics and Quality Assessment.

| Author | Year | Study Design | Year Range | Sample | Region<sup>¶</sup> | Patient Source | Follow-up | Rural/Urban Classification | Quality |
| --- | --- | --- | --- | --- | --- | --- | --- | --- | --- |
| Agrawal et al (14) | 2017 | Prospective single-centre cohort | 6 months | 337 | Asia | ED | Hospital Discharge | Not defined | Moderate |
| Andelic et al (16) | 2012 | Population-based prospective | 2009 – 2010 (2 yr) | 359 | Europe | Hospital | Hospital Discharge | Region-specific<sup>§</sup> | High |
| Asemota et al (26) | 2013 | Population-based retrospective | 2005 – 2009 (4 yr) | 139,798 | North America | Hospital | Hospital Discharge | Not defined | Moderate |
| Berry et al (5) | 2010 | Population-based retrospective | 2000 – 2006 (6 yr) | 95,485 | Oceania | Hospital | None | Formal Classification<sup>‡</sup> | Moderate |
| Brown et al (18) | 2019 | Population-based retrospective | 2008 – 2014 (7 yr) | NR | North America | Death registry | Death | Formal Classification | Moderate |
| Chan et al (43) | 2005 | Prospective multi-centre cross-sectional | 1998 – 2001 (4 yr) | 165 | Asia | ED | None | Region-specific | High |
| Chapital et al (20) | 2007 | Retrospective single-centre cohort | 2000 – 2004 (5 yr) | 3,447 | North America | Hospital | Hospital Discharge | Region-specific | High |
| Cheng et al (27) | 2017 | Population-based retrospective | 2006 – 2013 (8 yr) | 93,793 | North America | Death registry | Death | Formal Classification | Moderate |
| Cheng et al (45) | 2020 | Population-based retrospective | 1999 – 2017 (19 yr) | 99,796 | Asia | Death registry | Death | Region-specific | Moderate |
| Chiang et al (46) | 2006 | Population-based retrospective | 2001 – 2004 (3 yr) | 592 | Asia | Hospital | Hospital Discharge | Region-specific | Moderate |
| Chiu et al (19) | 2007 | Population-based retrospective | 2001 (1 yr) | 7,228 | Asia | Hospital/death certificates | Hospital Discharge | Region-specific | High |
| Daugherty et al (28) | 2021 | Population-based retrospective | 2016 – 2018 (3 yr) | 181,227 | North America | Death registry | Death | Formal Classification | Moderate |
| Feigin et al (38) | 2013 | Population-based prospective and retrospective | 2010 – 2011 (1 yr) | 1,369 | Oceania | All Community | None | Region-specific | High |
| Gabella et al (15) | 1997 | Population-based retrospective | 1991 – 1992 (2 yr) | 7,056 | North America | Hospital/death certificates | Hospital Discharge/Death | Formal Classification | High |
| Gontkovsky et al (29) | 2006 | Prospective single-centre cohort | 1998 – 2002 (3.5 yr) | 111 | North America | Rehabilitation | 1 yr | Goodall et al method<sup>63</sup> | High |
| Graves et al (30) | 2019 | Retrospective multi-centre cohort | 2007 – 2011 (5.5 yr) | 387,846 | North America | Hospital | 180 days | Region-specific | High |
| Halldorsson et al (44) | 2007 | Population-based prospective | 1992 – 1993 (1 yr) | 550 | Europe | Hospital/death registry | Death | Region-specific | High |
| Harradine et al (39) | 2004 | Prospective multi-centre longitudinal | 1999 – 2001 (2 yr) | 198 | Oceania | Rehabilitation | 18 mths | Formal Classification | High |
| Harrison et al (40) | 2012 | Population-based retrospective | 2000 – 2006 (6 yr) | 103,782 | Oceania | Hospital | Hospital Discharge | Formal Classification | High |
| Johnstone et al (31) | 2003 | Prospective single-centre longitudinal | 2 yr | 78 | North America | Rehabilitation | VR Completion | Formal Classification | Moderate |
| Karwat et al<sup>†</sup> (11, 49) | 2009 | Population-based retrospective | 1999 – 2002 (4 yr) | 265 | Europe | Hospital | Hospital Discharge/Death | Not defined | Moderate |
| Leonhard et al (32) | 2015 | Population-based retrospective | 2009 – 2012 (4 yr) | 2,794 | North America | Hospital | Death | Formal Classification | High |
| Maier et al (17) | 2014 | Retrospective multi-centre cross-sectional | 2005 – 2008 (R)<br>2003 – 2007 (U)<br>(3.5 yr) | 680 | Africa | Hospital | None | Region-specific | Moderate |
| Ponsford et al (6) | 2012 | Retrospective single-centre cohort | 1984 – 2006<br>(24 yr) | 959 | Oceania | Rehabilitation | 2 yr | Region-specific | High |
| Pozzato et al (21) | 2019 | Population-based retrospective | 2007<br>(1 yr) | 6,827 | Oceania | Hospital | Hospital Discharge | Formal Classification | Moderate |
| Ratliff et al (33) | 2021 | Population-based retrospective | 2008 – 2014<br>(7 yr) | 3,180 | North America | Death registry | Death | Formal Classification | Moderate |
| Reid et al (34) | 2001 | Population-based retrospective | 1993<br>(1 yr) | 977 | North America | Hospital/death certificates | Hospital Discharge/Death | Goldsmith et al method<sup>64</sup> | High |
| Ring et al (41) | 1986 | Retrospective multi-centre cohort | 1997 – 1978<br>(2 yr) | 991 | Oceania | Hospital | Hospital Discharge/Death | Region-specific | High |
| Robertson & McConnel (35) | 2011 | Retrospective single-centre cohort | 5 yr | 444 | North America | Hospital | Hospital Discharge | Formal Classification | Moderate |
| Schootman & Fuortes (36) | 2000 | Population-based retrospective | 1995 – 1997<br>(3 yr) | 4,300,000 | North America | Ambulatory care | End of Care Episode | Region-specific | Moderate |
| Simpson et al (42) | 2016 | Prospective, multi-centre cross-sectional | 2007 – 2008<br>(1 yr) | 503 | Oceania | Rehabilitation | 6 mths (minimum) | Formal Classification | High |
| Stewart et al (37) | 2014 | Retrospective single-centre cross-sectional | 2006 – 2011<br>(6 yr) | 2,112 | North America | ED | None | Region-specific | High |
| Tesfaw et al (47) | 2021 | Prospective single-centre cross-sectional | 2019<br>(2 mths) | 370 | Africa | Hospital | Hospital Discharge | Not defined | Moderate |
| Woodward et al (22) | 1984 | Population-based retrospective | 1980-1981<br>(2 yr) | 12,201 | Oceania | Hospital | Hospital Discharge | Formal Classification | Moderate |
| Yates et al (48) | 2006 | Retrospective single-centre cohort study | 1997 – 2003<br>(6 yr) | 11,700 | Europe | ED | None | Not defined | Moderate |

<sup>†</sup> Karwat et al (2009A and 2009B) report different outcomes on the same patient population, and are therefore considered as one study. <sup>‡</sup> Locations of Asian studies included India (14), Malaysia (43), China (45), and Taiwan (19, 46). European study locations were Norway (16), Iceland (44), Poland (11, 49), and the United Kingdom (48). Seven of the North American studies were national studies (18, 26-28, 30, 33, 36), with others state-based (Hawaii (20), Colorado (15), Mississippi (29), Missouri (31), Oregon (32), Minnesota (34), and Texas (35)), and one Canadian study (37). Oceania studies were all conducted in Australia with the exception of Feigin et al (38) which was a New Zealand study. The African studies were conducted in Tanzania (17) and Ethiopia (47). <sup>§</sup> Authors have designated a rural or urban label to a specific city, county or region based on its population, geographic or service provision characteristics. <sup>‡</sup> Authors have utilised a pre-existing nationally recognised method of classification. ED, emergency department; VR, Vocational Rehabilitation; R, rural; U, urban.

### Patient cohort

Seventeen studies included patients of all ages, while 12 reported on pediatric, adolescent, and/or young adult patients only (Table 2) (5, 26, 30, 32, 34, 35, 37, 40, 43, 44, 45, 46). Nine studies, comprising a total of 5,241 patients, included comparisons of patient age between rural and urban populations (6, 11, 16, 17, 31, 35, 37, 39, 43). As demonstrated in Figure 2, there was no significant difference in age at time of head injury between rural and urban patients (MD: 1.10, 95% CI [-3.17, 5.37, p=0.61). Males were over-represented in all cohorts, comprising 62-80% of head injury patients, with no difference between rural and urban areas (OR: 1.02, 95% CI [1.00, 1.04], p=0.11) (Table 2, Fig 3). A subgroup analysis also showed no statistically significant difference between the number of male patients in pediatric/adolescent or adult age groups between rural and urban populations (OR: 1.01, 95% CI [1.00, 1.03], p=0.13; and OR 1.16, 95% CI [0.82, 1.63], p=0.40, respectively) (Fig 3). Injuries of all severities were reported in 22 studies (5, 6, 14, 15, 18, 19, 20, 21, 26, 28, 29, 32, 33, 34, 36, 38, 40, 41, 44, 45, 46, 47), with the remaining studies reporting on either mild or severe traumatic head injury only (Table 2).

**Figure 2:**
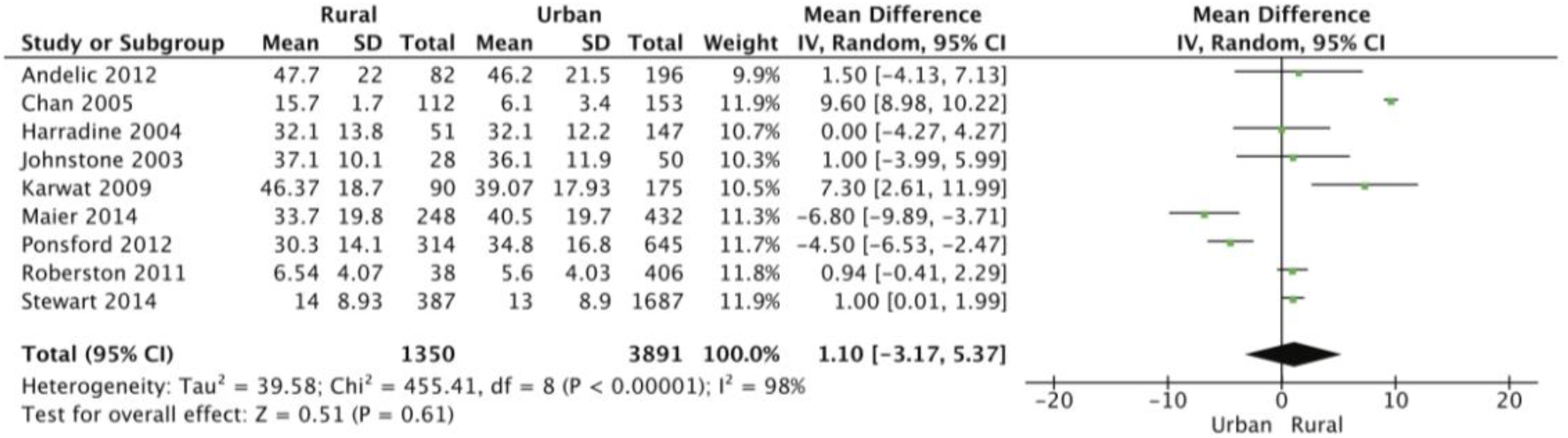
Forest plot demonstrating the mean difference in age (years) calculated using the random effects model. There was no statistical difference in age between rural and urban head trauma patients (MD: 1.10; 95% CI -3.17, 5.37; p=0.61). Mean [SD] for Stewart 2014 (37) was calculated using the methodology of Wan *et al* (25). SD, standard deviation; CI, confidence interval; I^2^, test of heterogeneity; MD, mean difference.

**Figure 3:**
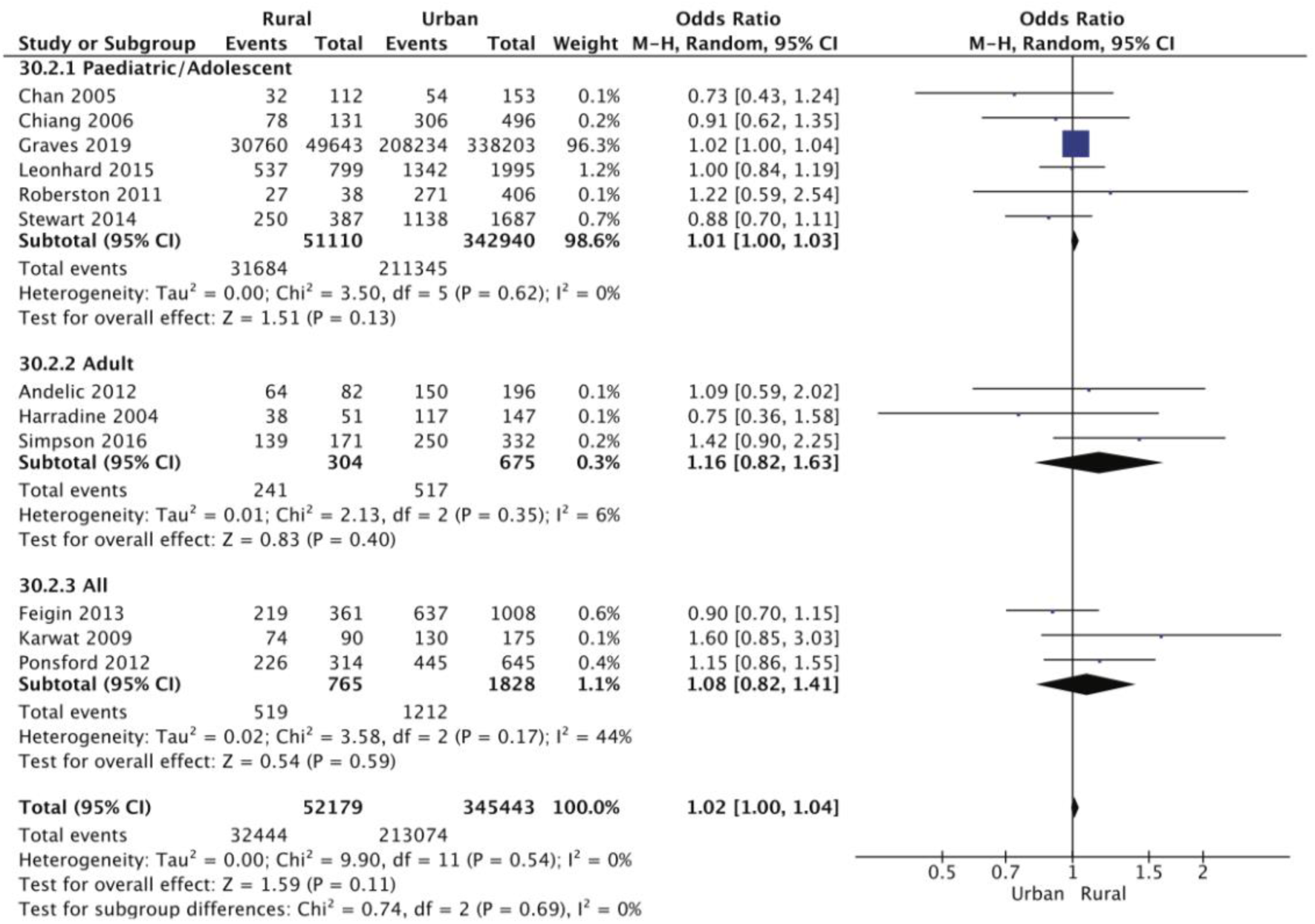
Forest plot demonstrating the odds ratio of male sex in rural and urban head trauma populations with subgroup analysis of paediatric/adolescent cohorts, adult cohorts, and studies incorporating all ages. The proportion of males suffering head trauma was comparable across rural and urban areas, regardless of age. SD, standard deviation; CI, confidence interval; I^2^, test of heterogeneity.

**TABLE 2:** Patient Characteristics.

| Author | Year | Age Group | Age Range (yr) | Male Sex (%) | Severity | Rural/Urban Ratio (%) |
| --- | --- | --- | --- | --- | --- | --- |
| Agrawal et al (14) | 2017 | All | 1-90 | 271 (80.4%) | All | 274 (72.6%)/54 (27.4%) |
| Andelic et al (16) | 2012 | Adult | >16 | 214 (77%) | Severe | 82 (29%)/196 (71%) |
| Asemota et al (26) | 2013 | Adolescent/Young Adult | 10-19 | 99,047 (71%) | All | 7,631 (5%)/132,167 (95%) |
| Berry et al (5) | 2010 | Pediatric | 0-14 | 61,179 (64%) | All | 11,160 (50.9%)/10,719 (50.0%) |
| Brown et al (18) | 2019 | All | NR | NR | All | NR |
| Chan et al (43) | 2005 | Pediatric/Adolescent | 2-18 | NR | Mild | 112 (42%)/153 (58%) |
| Chapital et al (20) | 2007 | All | 0-106 | 2573 (75%) | All | 358 (10.4%)/3,089 (89.6%) |
| Cheng et al (27) | 2017 | All | 0->75 | NR | NR | NR |
| Cheng et al (45) | 2020 | Adolescent/Young Adult | 0-19 | NR | All | NR |
| Chiang et al (46) | 2006 | Adolescents | 13-18 | 306 (65%) | All | 131 (22%)/469 (78%) |
| Chiu et al (19) | 2007 | All | NR | 4698 (65%) | All | 1,474 (20.4%)/5,754 (79.6%) |
| Daugherty et al (28) | 2021 | All | NR | NR | All | NR |
| Feigin et al (38) | 2013 | All | 0->65 | 856 (63%) | All | 361 (26.4%)/1,008 (73.6%) |
| Gabella et al (15) | 1997 | All | 0->65 | 4598 (67%) | All | 1,338 (19%)/5,525 (81%) |
| Gontkovsky et al (29) | 2006 | Adult | >16 | 79 (71%) | All | NR |
| Graves et al (30) | 2019 | Pediatric/Adolescent | 0-18 | 238,994 (62%) | Mild | 49,643 (13%)/338,203 (87%) |
| Halldorsson et al (44) | 2007 | Pediatric/Adolescent | 0-19 | NR | All | 141 (26%)/409 (74%) |
| Harradine et al (39) | 2004 | Adult | 16-65 | 155 (78%) | Severe | 51 (26%)/147 (74%) |
| Harrison et al (40) | 2012 | Adolescent/Young Adult | 15-24 | NR | All | 13,146 (47.2%)/14,728 (52.8%) |
| Johnstone et al (31) | 2003 | Adult | NR | 55 (71%) | NR | 28 (35.9%)/50 (64.1%) |
| Karwat et al<sup>†</sup> (11, 49) | 2009 | All | 0->65 | 204 (77%) | NR | 90 (34%)/175 (66%) |
| Leonhard et al (32) | 2015 | Pediatric/Adolescent | 0-19 | 1879 (67%) | All | 799 (28.6%)/1,995 (71.4%) |
| Maier et al (17) | 2014 | All | 0.2-100 | NR | NR | 248 (36.5%)/432 (63.5%) |
| Ponsford et al (6) | 2012 | All | 11-89 | 671 (70%) | All | 314 (32.7%)/645 (67.3%) |
| Pozzato et al (21) | 2019 | All | 0->70 | 2180 (74.5%) | All | 2,240 (33.3%)/4,482 (66.7%) |
| Ratliff et al (33) | 2021 | All | NR | NR | All | NR |
| Reid et al (34) | 2001 | Pediatric/Adolescent | 0-19 | NR | All | 343 (35.1%)/634 (64.9%) |
| Ring et al (41) | 1986 | All | 0->75 | 721 (73%) | All | 309 (31.2%)/682 (68.8%) |
| Robertson & McConnel (35) | 2011 | Pediatric/Adolescent | 0-18 | 298 (67%) | Severe | 38 (8.6%)/406 (91.4%) |
| Schootman & Fuortes (36) | 2000 | All | 0->75 | NR | All | NR |
| Simpson et al (42) | 2016 | Adult | 18-65 | 389 (77%) | Severe | 171 (34%)/332 (66%) |
| Stewart et al (37) | 2014 | Pediatric/Adolescent | 0-18 | 1415 (67%) | Mild | 387 (19%)/1,687 (81%) |
| Tesfaw et al (47) | 2021 | Adult | >18 | 265 (72%) | All | 259 (70%)/111 (30%) |
| Woodward et al (22) | 1984 | All | 0->75 | NR | NR | 3,971 (32.5%)/8,230 (67.5%) |
| Yates et al (48) | 2006 | All | 0->85 | NR | NR | NR |

<sup>†</sup> Karwat et al (2009A and 2009B) report different outcomes on the same patient population, and are therefore considered as one study. NR, not reported.

Population-adjusted overall incidence rate of traumatic head injury was reported in 12 studies (Table 3) (14, 16, 19, 22, 32, 34, 36, 37, 38, 40, 44, 48). The incidence rate per 100,000 was reported to be higher in rural populations in seven out of the 12 studies (14, 16, 19, 22, 32, 38, 40), with a trend for higher rates in more remote areas (Table 3) (14, 40).

**TABLE 3:** Incidence Rate of Head Trauma (/100,000 persons) in Rural and Urban Populations.

| Author | Year | Rural | Urban | p Value |
| --- | --- | --- | --- | --- |
| Andelic et al<sup>†</sup> (16) | 2012 | 5.9 (North)<br>4.3 (Central) | 5.0 (West)<br>4.1 (Southeast) | NR |
| Chiu et al (19) | 2007 | 417 | 218 | NR |
| Feigin et al<sup>‡</sup> (38) | 2013 | 73 | 31 | NR |
| Gabella et al<sup>†</sup> (15) | 1997 | 123.9 (Rural non-remote)<br>172.1 (Rural remote) | 97.8 (CMSA)<br>94.7 (Other metro) | NR |
| Halldorsson et al (44) | 2007 | 367 | 864 | NR |
| Harrison et al (40) | 2012 | 664.2 (Inner regional)<br>949.9 (Outer regional)<br>366.9 (Remote)<br>1680.2 (Very remote) | 522.1 (Major city) | NR |
| Leonhard et al (32) | 2015 | 107 | 71 (Large metropolitan)<br>59 (Small/medium metropolitan) | NR |
| Reid et al (34) | 2001 | 76.1 | 72.4 | 0.046 |
| Schootman & Fuortes (36) | 2000 | 410 | 570 | NR |
| Stewart et al (37) | 2014 | 220 | 350 | NR |
| Woodward et al (22) | 1984 | 570 | 430 | <0.001 |
| Yates et al (48) | 2006 | 223.8 (All HI)<br>29.6 (MSHI) | 826.9 (All HI)<br>55.6 (MSHI) | NR |

<sup>†</sup> Age-adjusted. <sup>‡</sup> Moderate-severe head injury. NR, not reported; CMSA, Consolidated metropolitan statistical area; HI, head injury; MSHI, moderate-severe head injury.

### Cause of injury

#### Transport-related head trauma

Traumatic head injuries caused by transport accidents were 1.3-fold more likely to occur in rural populations than in urban environments (p=0.001) (Fig 4). A subgroup analysis by age groups of 15 studies and 79,558 patients revealed that transport-related head injuries were significantly more common in rural pediatric and adolescent residents compared with their urban counterparts (OR: 1.27, 95% CI [1.10, 1.47], p<0.00001). Four studies reported on different vehicle types, which enabled a further analysis of the types of transport accidents across 10,526 patients (6, 19, 37, 43). The analysis revealed that there was no difference in car or bicycle accidents causing head injury between rural and urban groups (OR: 1.32, 95% CI [0.82, 2.07], p=0.026; and OR: 0.70, 95% CI [0.37, 1.30, p=0.26, respectively) (Fig 5AD). However, patients residing in urban locations were 64% more likely to suffer an injury as a pedestrian than those living rurally (OR: 0.36, 95% CI [0.17, 0.77], p=0.008) (Fig 5B), while head trauma resulting from accidents involving other motorized vehicles, such as all-terrain vehicles (ATVs) and motorcycles, was over 3.5 times more likely to have occurred in a rural setting (OR: 3.63, 95% CI [1.58, 8.35], p=0.002) (Fig 5C).

**Figure 4:**
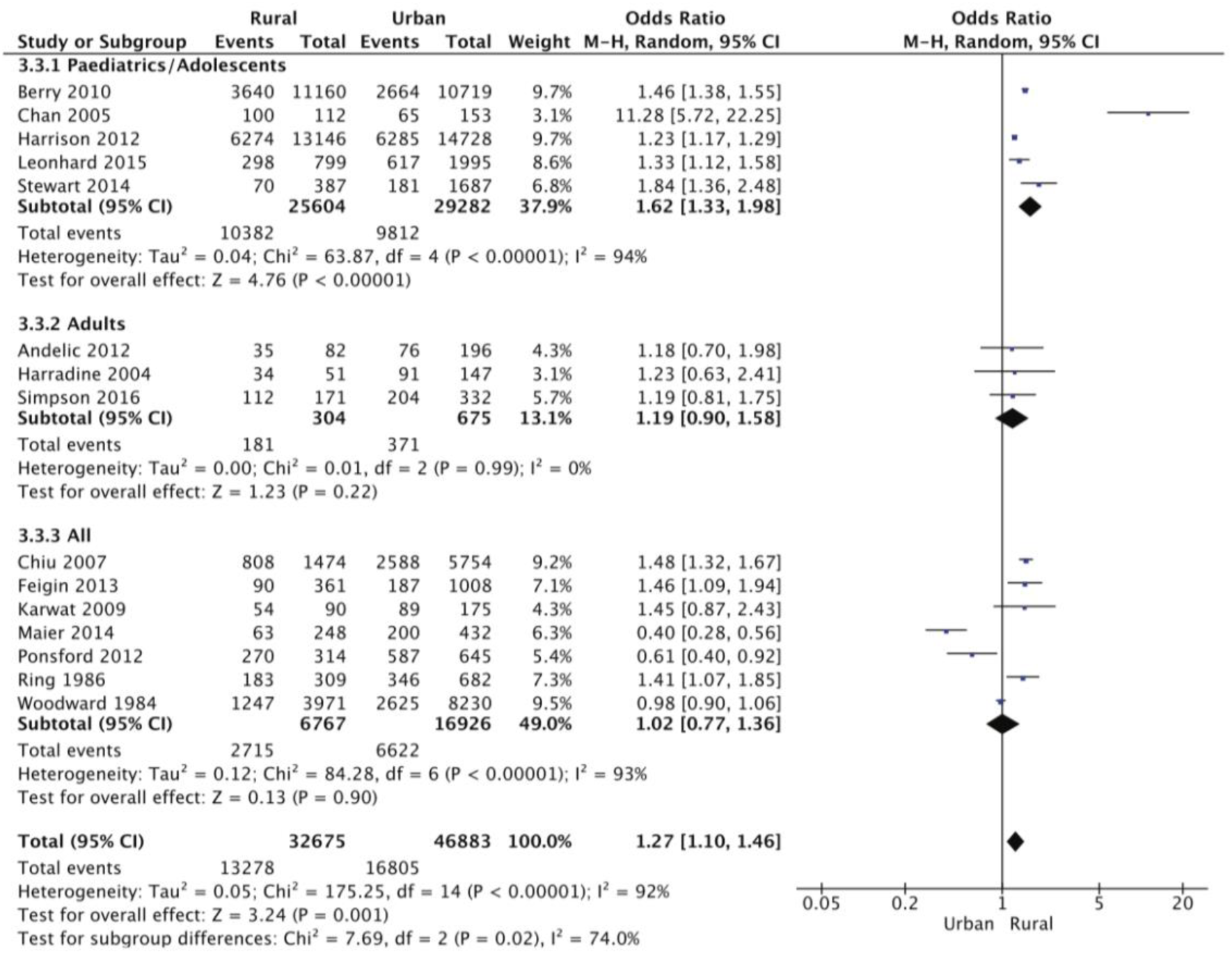
Forest plot demonstrating the odds ratio of transport being the cause of head trauma in rural and urban populations with subgroup analysis of pediatric/adolescent cohorts, adult cohorts, and studies incorporating all ages. Overall, rural residents were significantly more likely to suffer head trauma resulting from transport accidents, particularly those in pediatric and adolescent age groups (OR: 1.62; 95% CI 1.33, 1.98; p<0.00001). CI, confidence interval; I^2^, test of heterogeneity; OR, odds ratio.

**Figure 5:**
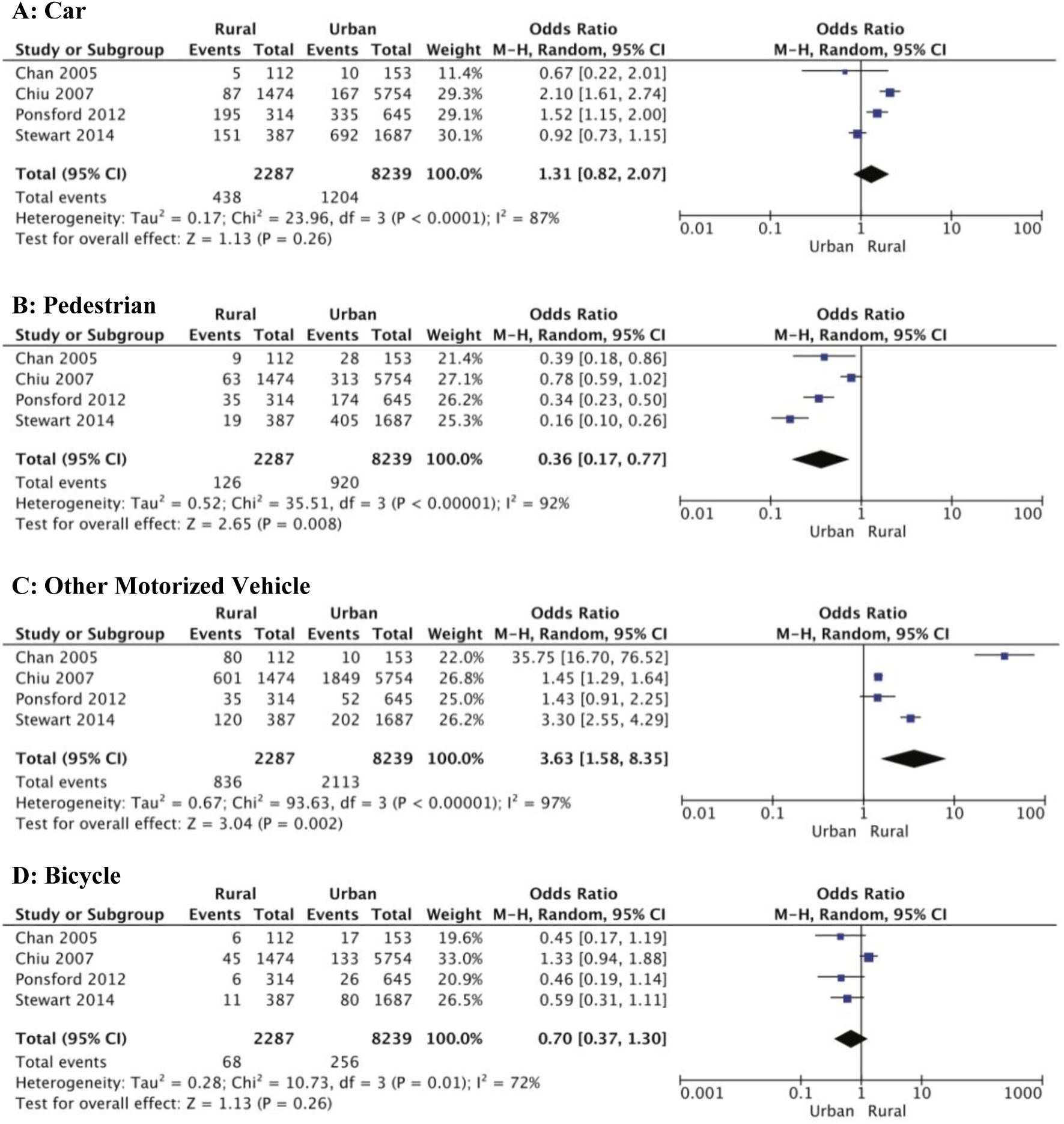
Forest plots showing types of transport accidents causing traumatic head injury in rural and urban populations. A, Car; B, Pedestrian; C, Other motorized vehicle; D, Bicycle. Pedestrian-related head injuries were 64% more likely in urban environments (p=0.008), whereas motorized vehicles such as all-terrain vehicles, quad bikes, and motorcycles, were responsible for more injuries in rural areas (OR: 3.63; 95% CI 1.58, 8.35; p=0.002). CI, confidence interval; I^2^, test of heterogeneity; OR, odds ratio.

#### Other causes of head trauma

A total of 79,478 patients from 15 studies were analyzed for fall-related head injury (5, 6, 16, 19, 22, 32, 37, 38, 39, 40, 41, 42, 43, 46, 49). Urban residents were 27% more likely to sustain head injury following a fall when compared to those from rural areas (OR: 0.73, 95% CI [0.66, 0.81], p<0.00001) (Fig 6). Subgroup analysis by age group shows a particularly high burden of fall-related head trauma in children and adolescents in urban settings (OR: 0.65, 95% CI [0.52, 0.81], p=0.0002).

**Figure 6:**
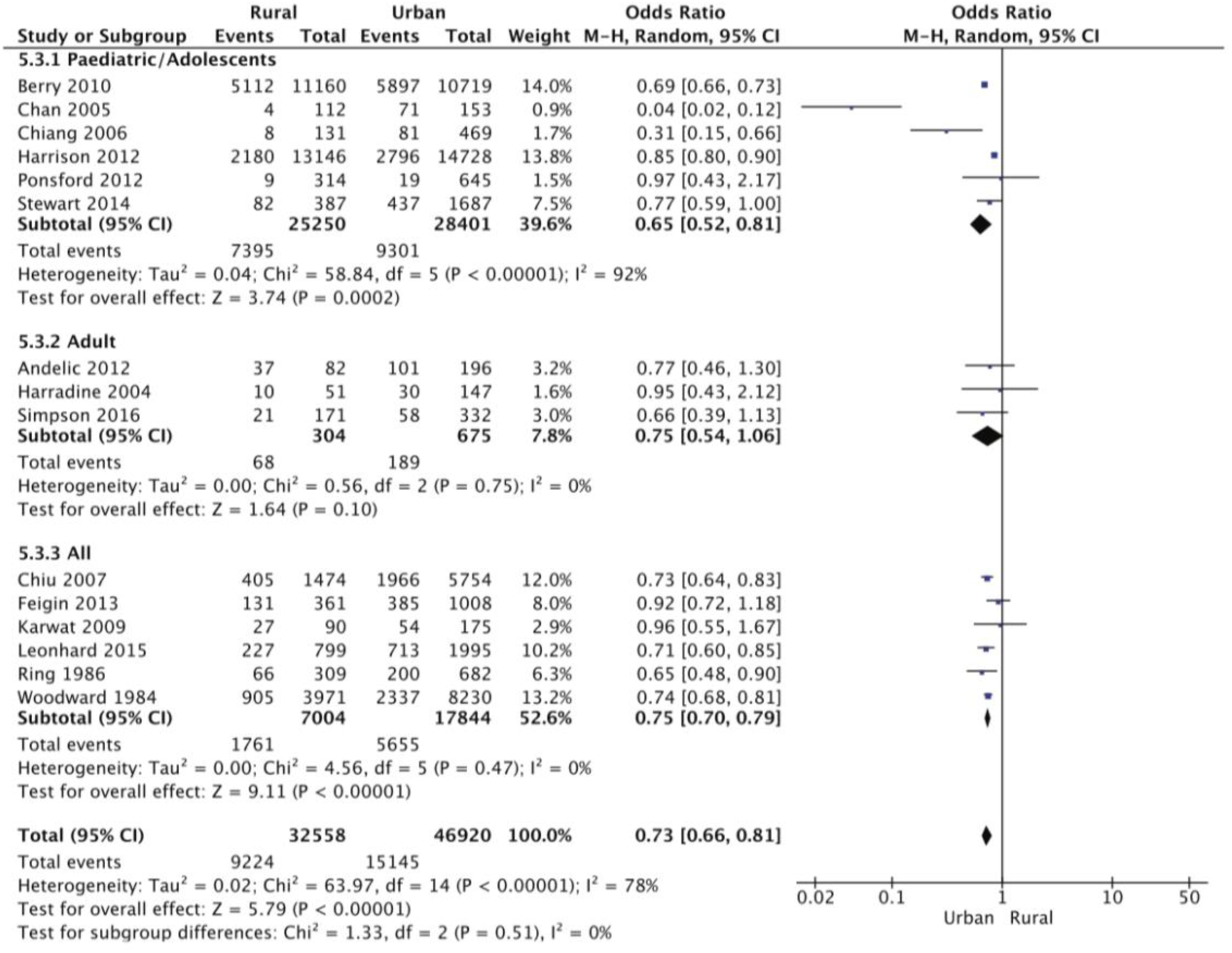
Forest plot demonstrating the odds ratio of falls being the cause of head trauma in rural and urban populations with subgroup analysis of pediatric/adolescent cohorts, adult cohorts, and studies incorporating all ages. Overall, urban residents were 27% more likely to suffer head trauma resulting from falls (p<0.00001). The odds of fall-induced head trauma was significantly less in rural children and adolescents (OR: 0.65; 95% CI 0.52, 0.81; p<0.0002). CI, confidence interval; I^2^, test of heterogeneity; OR, odds ratio.

Assault was investigated as a cause of head trauma in rural and urban populations in 11 studies, representing a total 73,699 patients (5, 6, 16, 17, 19, 22, 38, 39, 40, 42, 43, 49). As shown in Figure 7, no difference was found between urban and rural patients (OR: 0.84, 95% CI [0.59, 1.18], p=0.30). However, when accounting for injury severity, assault-related mild head injury was approximately half as likely to occur rurally (OR: 0.52, 95% CI [0.29, 0.94], p=0.03) (Fig 7). There was no difference in sports injuries or other causes of injury such as work accidents, exposure to animate and inanimate mechanical forces, or use of firearms, between rural and urban populations (OR: 0.99, 95% CI [0.59, 1.65], p=0.97; and OR: 1.18, 95% CI [0.77, 1.81], p=0.44, respectively) (Fig 8AB).

**Figure 7:**
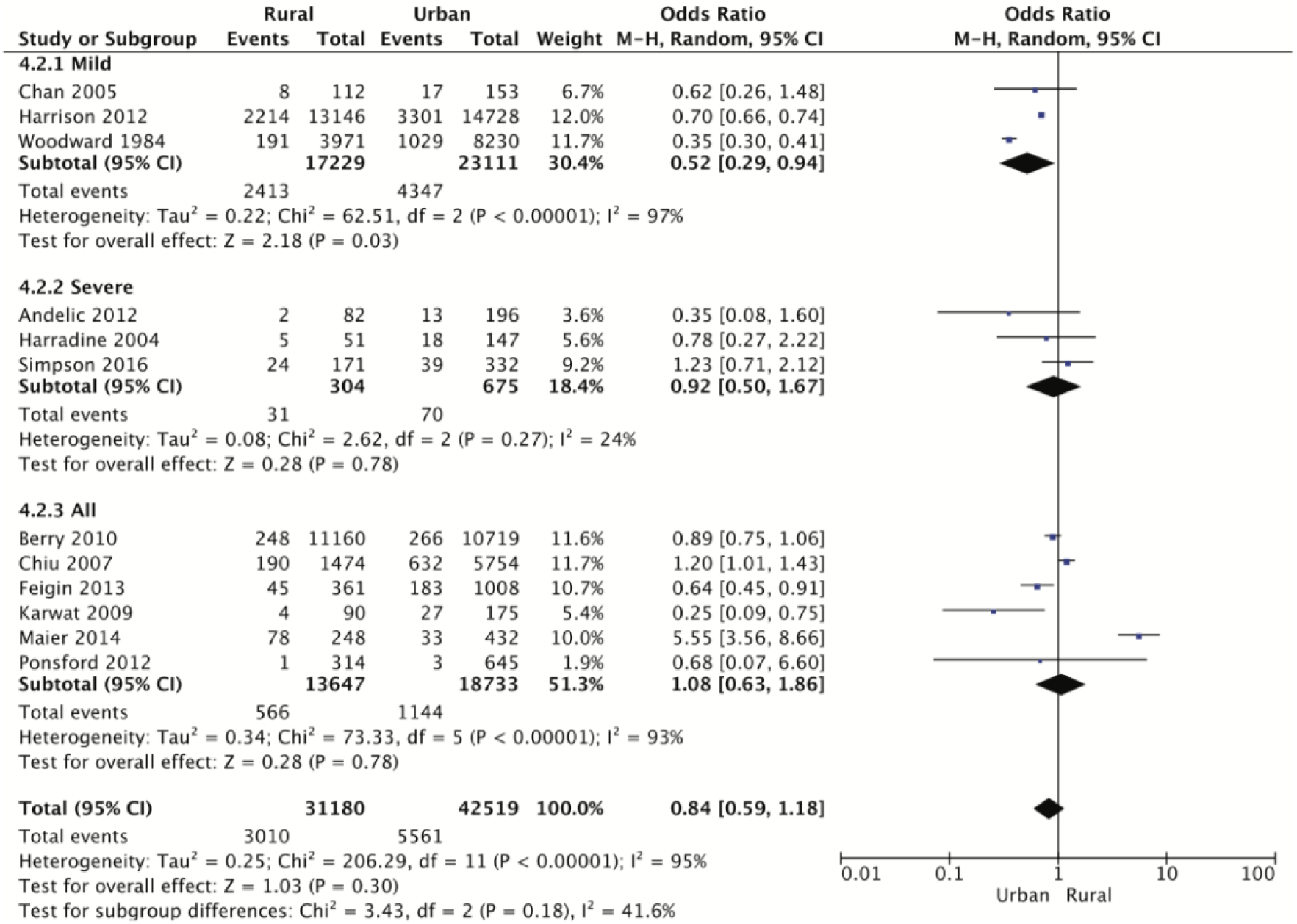
Forest plot demonstrating the odds ratio of assault being the cause of head trauma in rural and urban populations with subgroup analysis of mild, severe, and injury severities. Overall, assault-related head injury was comparable across rural and urban populations, except mild head injuries caused by assault which were ∼50% less likely in rural areas (p=0.03). CI, confidence interval; I^2^, test of heterogeneity.

**Figure 8:**
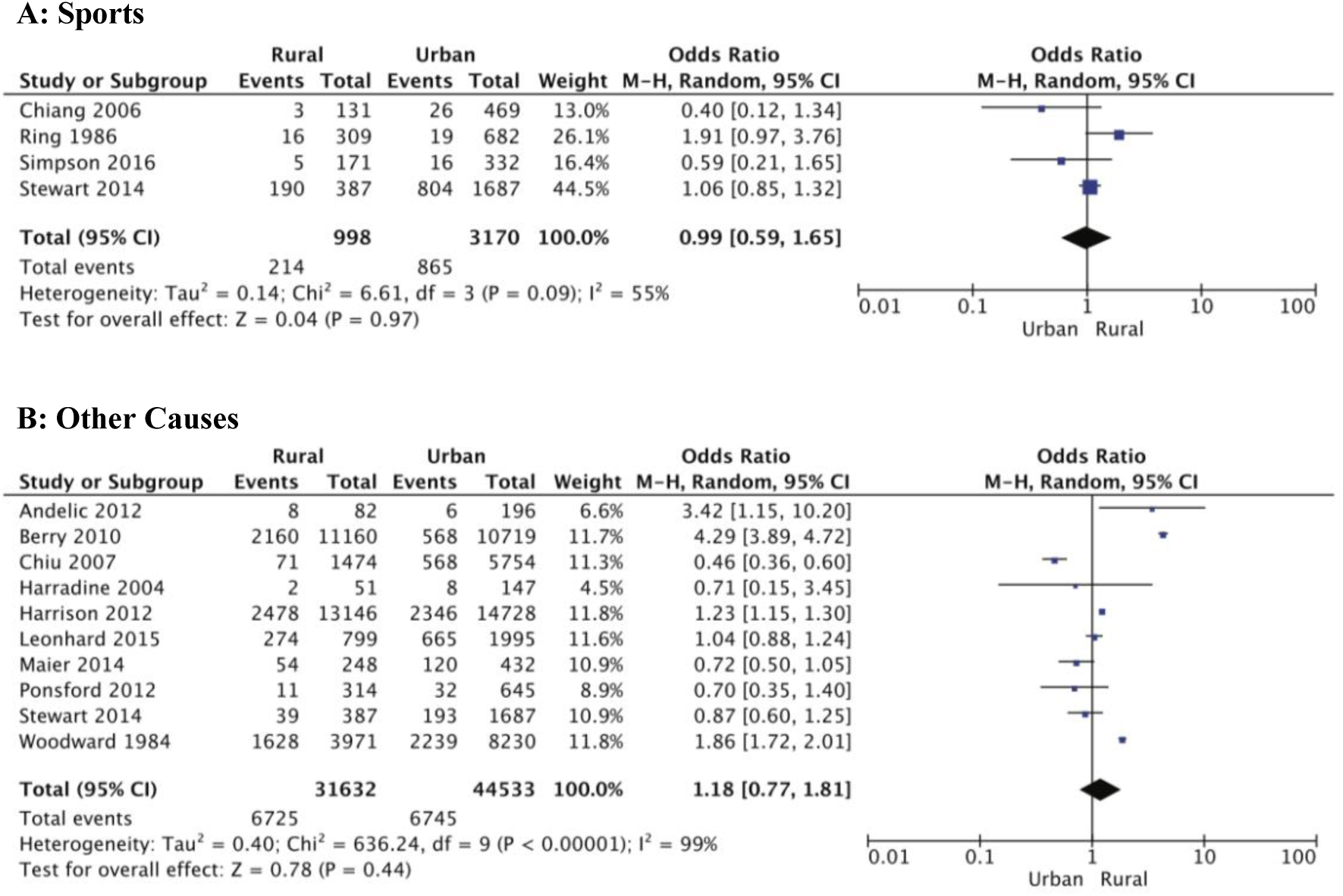
Forest plots showing no difference in (A) sports-related and (B) other causes of head trauma in rural and urban populations. Other causes included exposure to animal and inanimate mechanical forces, work-related accidents, and use of firearms. CI, confidence interval; I^2^, test of heterogeneity.

### Injury severity

Rates of severe traumatic head injury between rural and urban populations were reported by five studies, together representing 14,399 patients (19, 21, 32, 34, 46). Rural patients were 28% more likely to have obtained a severe traumatic head injury than urban patients (OR: 1.28, 95% CI [1.04, 1.58], p=0.02) (Fig 9A). However, the mean Glasgow Coma Scale (GCS), which was measured in 1,879 patients from four studies (6, 16, 35, 39), did not show a statistically significant difference between rural and urban populations (MD: -0.25, 95% CI [-0.82, 0.34], p=0.39) (Fig 9B). Rural patients were almost half as likely to have a normal CT following traumatic head injury when compared to urban patients (OR: 0.52, 95% CI [0.41, 0.67], p<0.00001) (Fig 9C). In contrast, the presence of skull fractures and intracranial hemorrhage was comparable across both populations (OR: 0.71, 95% CI [0.17, 2.96], p=0.64; and OR: 1.05, 95% CI [0.68, 1.63], p=0.82, respectively) (Fig 9DE).

**Figure 9:**
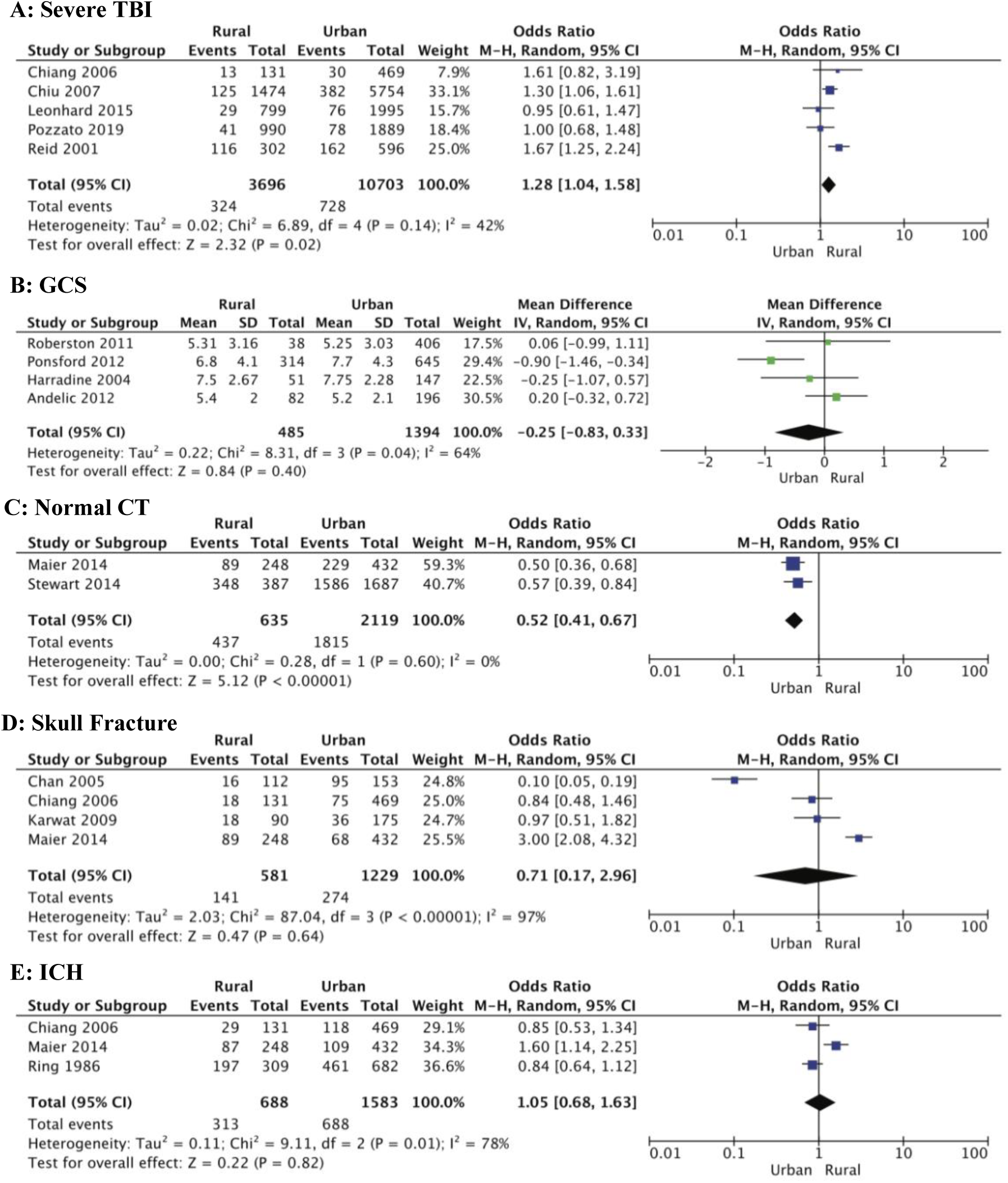
Forest plots showing measures of head injury severity in rural and urban populations. A, Proportion of severe head injury (GCS 3-8); B, GCS (mean [SD); C, Normal CT findings; D, CT-diagnosed skull fracture; E, CT-diagnosed ICH. Rural residents were significantly more likely to suffer severe head trauma (OR: 1.28; 95% CI 1.04, 1.58; p=0.02), and less likely to have a normal CT (OR: 0.52; 95% CI 0.41, 0.67; p<0.00001). Mean [SD] GCS for Harradine 2004 (39) was calculated using the methodology of Wan *et al* (25). GCS, Glasgow Coma Scale; CT, computed tomography; ICH, intracranial hemorrhage; CI, confidence interval; I^2^, test of heterogeneity; OR, odds ratio.

### Clinical symptoms

Various signs and symptoms associated with traumatic head injury were investigated by these studies. A total of 607 rural and urban patients were reported to have experienced loss of consciousness (LOC) and/or altered level of consciousness (ALOC) across three different studies (17, 37, 43). Meta-analysis revealed a significant disparity between rural and urban patients, with a 5-fold increased risk in the rural population (OR: 5.04, 95% CI [1.08, 23.62], p=0.04) (Fig 10A). Other clinical symptoms, including headache, seizures, and nausea and vomiting, did not differ between rural and urban patients (Fig 10B-D).

**Figure 10:**
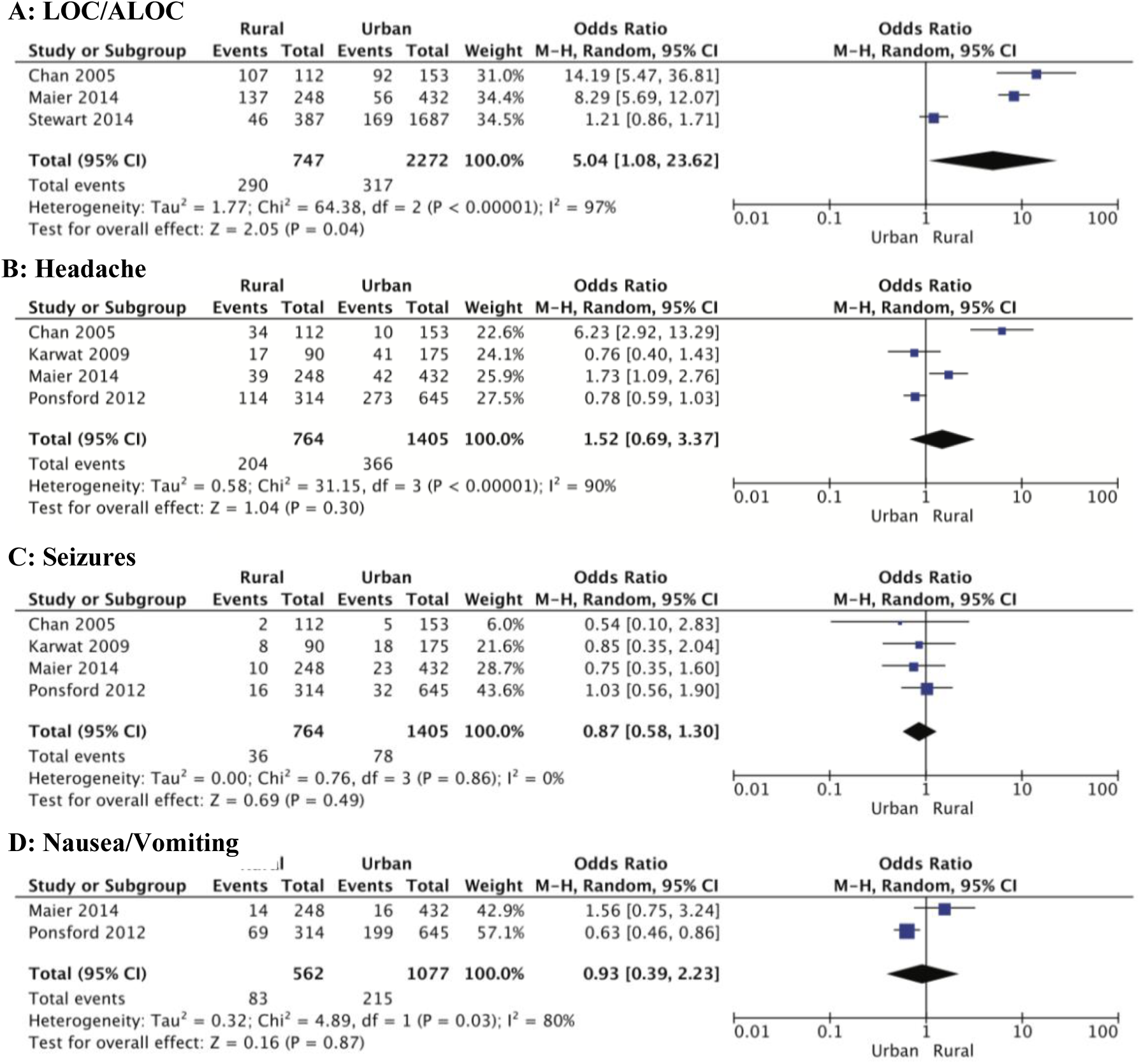
Forest plots showing clinical symptoms after head injury in rural and urban populations. A, LOC/ALOC; B, Headache; C, Seizures; D, Nausea/Vomiting. Rural residents were five-fold more likely to suffer loss of, or altered, consciousness (p=0.04). Incidence of headache, seizures, and nausea or vomiting was similar in rural and urban patients. LOC, loss of consciousness; ALOC, altered level of consciousness; CI, confidence interval; I^2^, test of heterogeneity.

### Outcomes

#### Mortality

Mortality was the major outcome measure reported in 16 studies (14, 15, 16, 18, 19, 20, 21, 22, 27, 32, 33, 34, 35, 41, 46, 49). Population-adjusted mortality rates and rate ratios were significantly higher in all rural cohorts, with further increases as level of rurality increased (Table 4) (14, 32, 33). However, meta-analysis of 11 studies including 32,984 patients which reported mortality events, failed to show a statistical difference between rural and urban populations (OR: 1.09, 95% CI [0.73, 1.61], p=0.67) (Fig 11A). Sensitivity analysis involving removal of the Woodward *et al* (22) and Pozzato *et al* (21) cohorts significantly reduced heterogeneity (I^2^ 86% to 29%), and supported the mortality rate and rate ratio data (Table 4), with a 1.5-fold increased risk of mortality in rural patients (OR: 1.49, 95% CI [1.21, 1.84), p=0.00002). There was no evidence of significant publication bias as indicated by the largely symmetrical funnel plot (Fig 11B).

**Figure 11:**
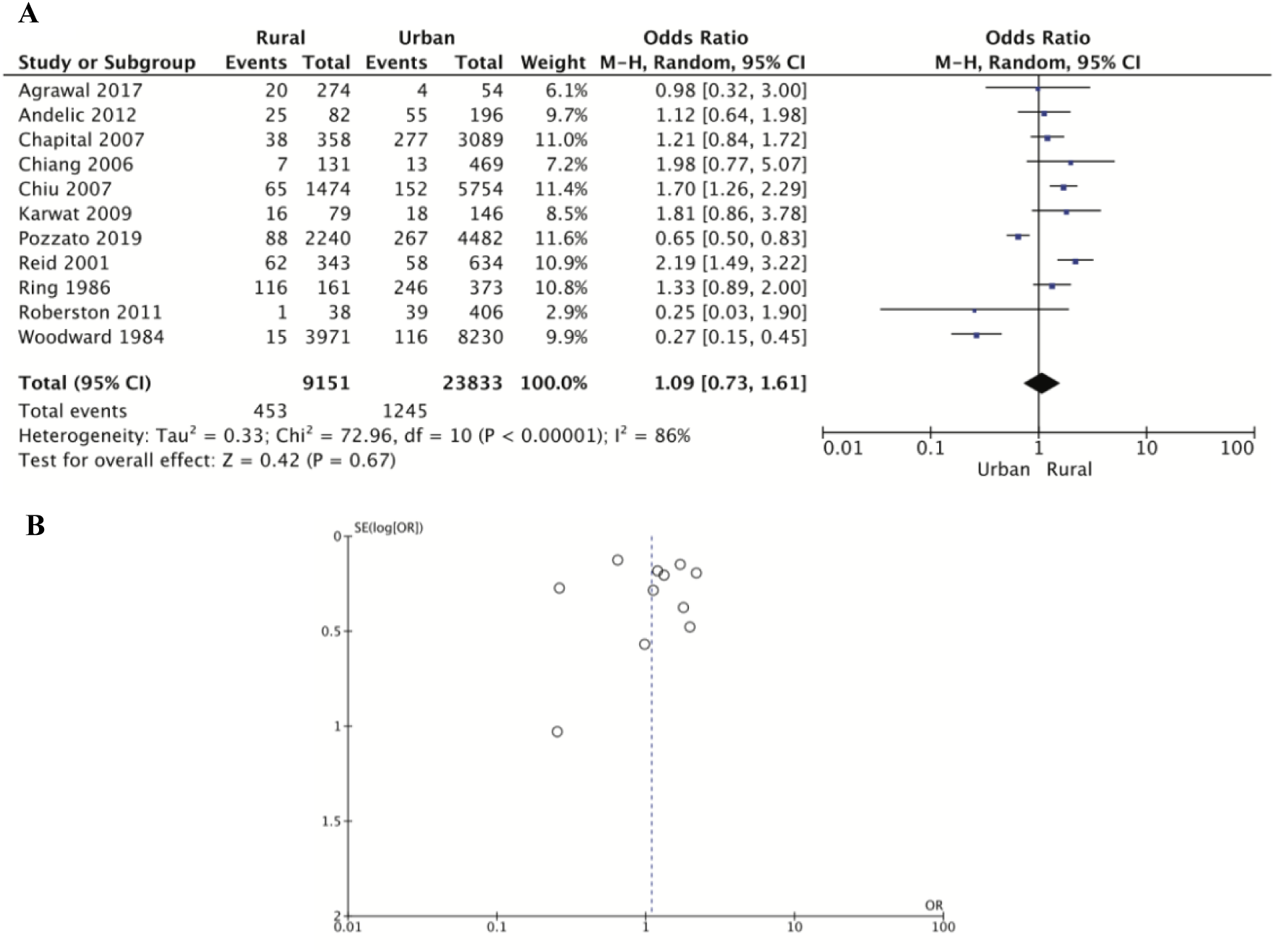
A, Forest plot showing mortality incidence in rural and urban head trauma populations calculated using the random effects model. Mortality was comparable across rural and urban areas. B, Funnel plot of publication bias. CI, confidence interval; I^2^, test of heterogeneity.

**TABLE 4:**
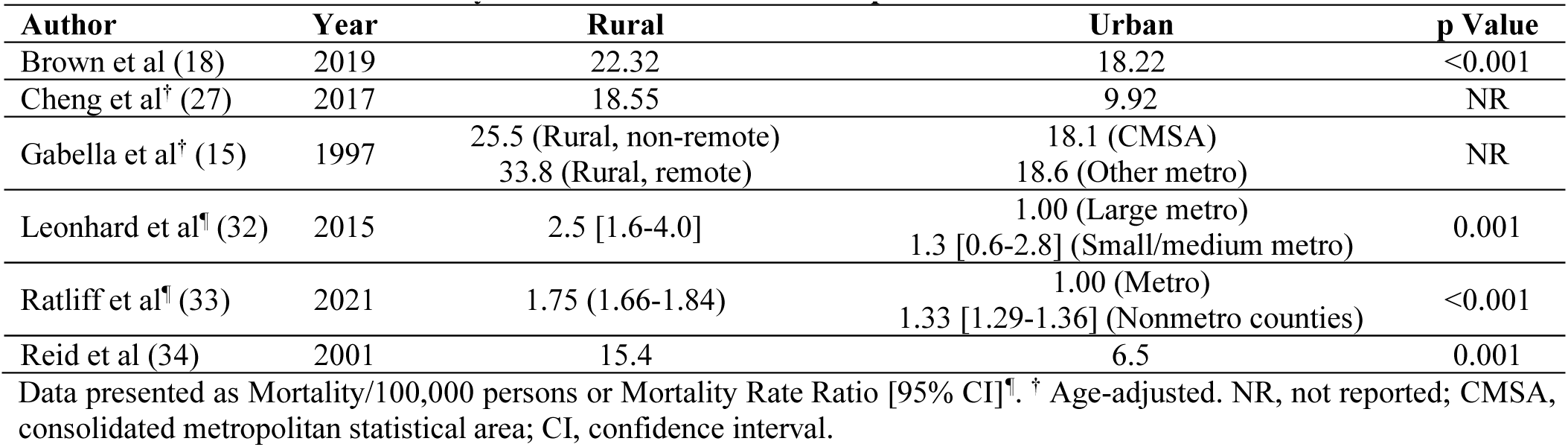
Head Trauma Mortality Rate in Rural and Urban Populations.

#### Other outcomes

Discharge status of head trauma patients was reported in three studies, comprising 22,103 patients (Fig 12A) (19, 22, 46). The odds of rural patients suffering severe disability or being in a vegetative state on hospital discharge was not significantly greater when compared to urban patients (OR: 1.42, 95% CI [0.44, 4.62], p=0.56). However, having a good recovery following traumatic head injury was significantly more likely in urban compared with rural residents (OR: 0.53, 95% CI [0.35, 0.81], p=0.003) (Fig 12B). The average length of hospital stay (LOS) for head trauma care was several days shorter for urban patients, however this difference did not reach statistical significance (MD -3.23, 95% CI [-10.08, 3.63], p=0.36) (Fig 12C). Finally, there were no differences in the likelihood of post-injury employment between rural and urban residents (OR: 1.06, 95% CI [0.59, 1.89], p=0.85) (Fig 12D).

**Figure 12:**
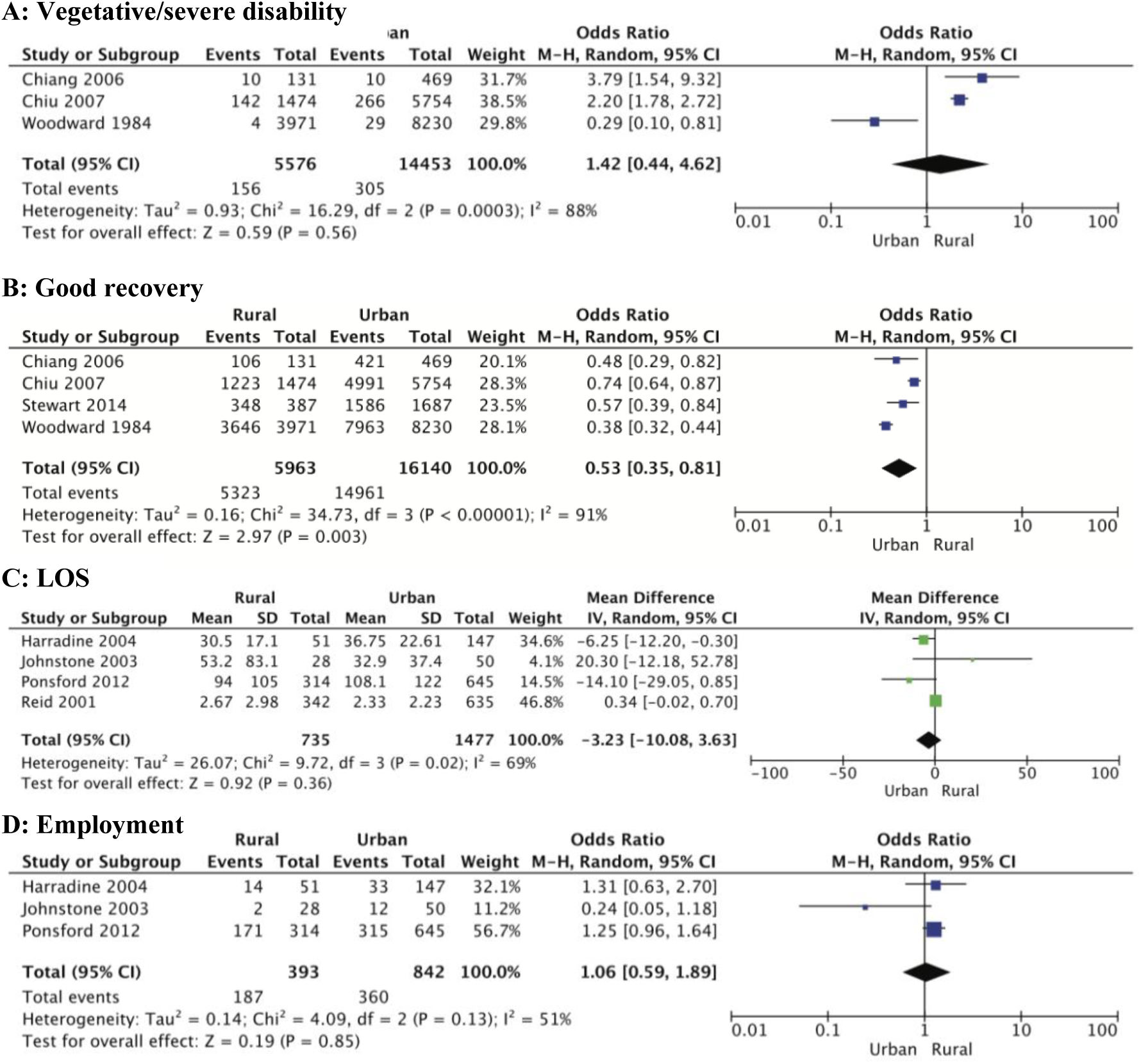
Forest plots showing outcomes after head injury in rural and urban populations. A, Vegetative/severe disability; B, Good recovery; C, Length of hospital stay (days); D, Employed post-injury. Urban residents were more likely to have a good recovery (p=0.003), and reduced hospital stay, however this difference was not statistically significant (MD: -3.23; 95% CI -10.08, 3.63; p=0.36). Mean [SD] LOS for Harradine 2004 (39) and Reid 2001 (34) was calculated using the methodology of Wan *et al* (25). LOS, length of stay; CI, confidence interval; I^2^, test of heterogeneity; MD, mean difference.

## Discussion

Head trauma is a global healthcare problem affecting up to 69 million people annually (1). It is a leading cause of morbidity and mortality, and successful treatment requires a time-critical approach (2, 50). Patients in rural areas may be disadvantaged by limited access to acute trauma care, complicated by longer transport times and distances to definitive care (51, 52). We conducted a systematic review and meta-analysis of 36 studies with approximately 2.5 million head trauma patients to address this question and report the following: First, the incidence of traumatic head injury was higher in males, regardless of location. Second, overall prevalence of head trauma was significantly higher in rural populations and involved more transport accidents compared to urban environments. Third, urban traumatic head injury patients were twice as likely to be discharged with a good outcome. These results will now be discussed.

### Head trauma is more prevalent in males regardless of geographical location

Our systematic review and meta-analysis showed a higher prevalence of head trauma in males than in females in both rural and urban environments (Table 2). We found males represented 62-80% of head injury patients in all cohorts, and there was no significant difference among pediatric/adolescent or adult age groups (Table 2, Fig 3). Similar to other types of traumatic injury, a higher incidence of head trauma in males is most likely related to an increased likelihood of involvement in high-risk activities, physical altercations, military service and contact sports (11, 17, 53).

### The incidence and severity of traumatic head injury is higher in rural areas

This meta-analysis confirmed a higher incidence of head trauma in rural settings compared to urban environments, and further showed that head injuries sustained in rural communities were more severe, indicated by the significantly higher proportion of rural patients with a GCS of 3-8 (Table 3, Fig 9A). Interestingly, a higher severity of injury in rural patients was not reflected in the mean GCS value, which did not significantly differ between rural and urban cohorts (Fig 9B). There are several possible reasons for this disparity. First is the fluctuating nature of GCS observed early after injury (54). Second are factors unrelated to the primary injury, such as the presence of alcohol, drugs, sedation and other medications (55, 56), and third, the challenge of accurate measurement of GCS in children, linguistically diverse people, and people with disabilities or cognitive deficits (38, 44, 54, 55). Furthermore, most of the published studies examined in the present analysis did not include prehospital data, and relied on the hospital admission GCS, which may not be an accurate representation of the true injury severity, particularly in patients with prolonged prehospital transport times.

An alternative to GCS and a more objective assessment of injury severity is non-invasive diagnostic imaging, such as CT. We found non-pathological (normal) CT was significantly more common amongst urban patients (Fig 9C), supporting our previous finding of fewer severe injuries in urban settings. However, this interpretation may be influenced by the greater availability and access to imaging modalities in urban hospitals. Rural hospitals may only perform imaging on more severely injured patients, reducing the number of CTs performed, and therefore the number of normal CT findings recorded (17).

A higher incidence of severe head trauma in rural locations was also supported by the proportion of patients experiencing loss of, or altered, consciousness after head injury, which was five times more prevalent in the rural patients (Fig 10A). Occurrence of other clinical symptoms, such as headache, nausea and vomiting, and seizures, was similar between rural and urban patients, however these are all non-specific and highly subjective, and susceptible to variable reporting by both patients and healthcare providers. They may also be influenced by medications, comorbidities, transport, and stress. Therefore, these symptoms may be significant when targeting supportive management on scene but provide minimal insight when determining severity.

### Head trauma due to transport accidents disproportionately affects young rural residents

Another major finding of our meta-analysis was that transport-related traumatic head injuries were 27% more likely to occur in rural locations (Fig 4). This risk more than doubled for children and adolescents. Head trauma resulting from motorized vehicle accidents (e.g., motorcycles and ATVs) was significantly more common in rural areas (Fig 5C). Several behavioural factors have been proposed as potential contributors to these findings. Cheng *et al* (2017) (27), Karwat *et al* (49) and Tesfaw *et al* (47) suggested lower rates of law-abiding behaviour and higher rates of risk-taking behaviour, including driving under the influence of alcohol, rurally. Greater occupational and recreational ATV use associated with rural farming regions may also account for the higher rate of head injuries resulting from motorized vehicle accidents in rural areas. However, there has been minimal discussion on the environmental factors associated with rural roads. The Centre for Accident Research and Road Safety Queensland have stated that the risk of sustaining a road crash injury increases with the degree of remoteness, with lower rates of safe driving practices contributing to this (57). However, rural areas also have several unique characteristics that predispose drivers to accidents, including lower road quality, unpredictable weather conditions, livestock and wildlife (58). These environmental factors, in addition to human factors, are likely somewhat responsible for the increased risk of transport-related head trauma in rural environments.

### Fall-related head trauma is more common in urban populations

Another interesting finding of our meta-analysis was the 2.7-fold greater odds of sustaining a traumatic head injury after a fall in urban populations, with children and adolescents from these areas having almost double the risk of their rural counterparts (Fig 6). Similarly, urban cohorts were twice as likely to sustain a mild head injury due to an assault, despite rural residents being more likely to experience intimate partner violence, which is a common cause of head trauma (59). The increased incidence of both fall-related head trauma, and assault-related mild head injury, may reflect greater health-seeking behaviours and access to healthcare in urban areas, and thereby increased reporting of these injuries.

### Mortality from head trauma is similar, but good recoveries are more likely in urban patients

Despite showing population-adjusted mortality rates and rate ratios were significantly higher in all rural cohorts, with further increases as level of rurality increased (Table 4, Fig 9A), our meta-analysis involving 11 studies including 32,984 patients showed no statistically significant difference in mortality incidence between rural and urban populations (Fig 11A). Importantly of the studies examined, Gabella *et al* (14) and Reid *et al* (34) were the only studies that specifically reported prehospital mortality. Previous research has established that shorter prehospital times are associated with improved head trauma survival (22, 60), and since rural patients have longer travel times and distances, it is possible they experienced greater prehospital mortality. However, unfortunately these prehospital statistics were not documented in the majority of studies of our meta-analysis. In addition, mortality due to head trauma can occur after hospital discharge and the follow-up time in some studies may not have been sufficient to capture all potential deaths and other complications (30).

Consistent with reduced severity and more normal CT findings, urban patients were 47% more likely to be discharged with a good outcome compared to their rural counterparts (Fig 12B). The initial analysis revealed no significant difference in severe disability or vegetative states between the two populations, however, a sensitivity analysis involving the exclusion of the Woodward *et al* study (22) revealed that rural residents were more than twice as likely to be discharged from hospital in a vegetative state or with severe disability (Fig 12A). This difference might be related to the standard of prehospital care when the study was conducted in 1984, reducing the chance of traumatic head injury patients even surviving to hospital discharge to be accounted for in this analysis. Moreover, the Woodward *et al* data (22) includes patients discharged to nursing homes, and admission to this type of facility may not accurately reflect the presence of severe head trauma as much as it reflects the multifactorial need for round-the-clock care.

### Strengths and limitations

To the best of our knowledge, this is the first time that a quantitative meta-analysis has been conducted to investigate epidemiological and outcome differences between rural and urban traumatic head injury patients. This comprehensive meta-analysis has included 36 studies from 14 countries with data spanning over 40 years. Most of these studies are population-based, which is considered the ideal approach to obtain objective measures and to understand disease patterns (21). Nevertheless, there are a number of limitations inherent in all head trauma research that must be considered. Studies of head injuries typically have considerable heterogeneity due to differences in defining and classifying head trauma, its severity and outcomes (21, 38). Of the 25 analyses performed in this study, 21 (84%) demonstrated substantial heterogeneity, indicated by high I^2^ values. In addition, the studies also varied in their classification of rurality, with some using population measurements whilst others used distance to a specific healthcare facility (Table 1, Appendix 7). Inclusion and exclusion criteria also differed across studies (Appendix 6). We mitigated the impact of heterogeneity as best as possible by using a random effects model in our meta-analysis.

Another limitation of all head trauma research is that mild injuries are frequently underreported. Traumatic head injury is referred to as a silent epidemic because mild injuries can present with few symptoms or sequelae and consequently, patients may not present to a healthcare facility, or are exclusively treated in outpatient settings (26, 30, 38). Therefore, research sourcing patients exclusively from hospitals or death registries, which was the case for 95% of the studies included in this meta-analysis, will potentially underreport mild cases. This issue is amplified when considering the barriers to accessing healthcare in rural areas. Therefore, it is likely that the true incidence of traumatic head injury, particularly in rural areas, is underestimated and skewed to higher severities.

Lastly, we have found that rural head trauma research lacks Indigenous representation in its datasets, preventing important subgroup analysis. Only five studies reported on Indigenous status in their patient cohorts (21, 26, 32, 38, 42), however, none evaluated rural/urban differences in this specific population. This is a concern because not only do Indigenous Peoples often represent a greater proportion of rural communities than urban, but they also experience significant health disparities as a result. In Australia, First Nations people have, on average, a life expectancy 10 years shorter than non-Indigenous Australians, and 15 years less if they live in remote or very remote areas (51, 61). Previous studies have shown Indigenous Australians and Americans are disproportionately affected by head trauma (9, 12, 62), however, the specific contribution of rurality has not been assessed. Therefore, it is critical to define the impact that ethnicity may have as a risk factor for traumatic head injury, particularly to develop acute care guidelines and preventative interventions.

### Recommendations for future research

Despite these limitations, ongoing epidemiological head trauma research is vital for identifying potential targets for all levels of prevention and management. We encourage the trauma research community to utilise standard definitions and classifications for head injury diagnosis, severity, and outcomes. Mortality alone should not be the only outcome measure reported because it does not consider the significant disability burden among survivors. Additionally, we recommend the inclusion of community, primary care, and other prehospital data, with adequate follow-up data. Finally, we strongly recommend the identification and subgroup analysis of Indigenous patients in datasets, and the use of population-based classifications for rurality. This will more accurately define rural/urban disparities associated with traumatic head injury and facilitate the development of evidence-based targeted interventions.

## Conclusions

Rurality is associated with greater incidence, severity, and poorer outcomes of traumatic head injury. Transport accidents are a significant cause of head trauma in rural environments. Future research should include primary care and prehospital data, as well as adequate follow-up for accurate incidence and mortality rates. The use of standardized severity and rural classifications, as well as the inclusion of Indigenous subgroup analyses are highly encouraged in future head trauma studies.

## Data Availability

All data produced in the present work are contained in the manuscript.

## Data Availability Statement

This systematic review and meta-analysis is a synthesis of existing published data, openly available in cited references.

## APPENDIX 1: PRISMA Checklist

| Section/topic | # | Checklist item | Reported on page # |
| --- | --- | --- | --- |
| **TITLE** |  |  |  |
| Title | 1 | Identify the report as a systematic review, meta-analysis, or both. | 1 |
| **ABSTRACT** |  |  |  |
| Structured summary | 2 | Provide a structured summary including, as applicable: background; objectives; data sources; study eligibility criteria, participants, and interventions; study appraisal and synthesis methods; results; limitations; conclusions and implications of key findings; systematic review registration number. | 2-3 |
| **INTRODUCTION** |  |  |  |
| Rationale | 3 | Describe the rationale for the review in the context of what is already known. | 4 |
| Objectives | 4 | Provide an explicit statement of questions being addressed with reference to participants, interventions, comparisons, outcomes, and study design (PICOS). | 4 |
| **METHODS** |  |  |  |
| Protocol and registration | 5 | Indicate if a review protocol exists, if and where it can be accessed (e.g., Web address), and, if available, provide registration information including registration number. | 5 |
| Eligibility criteria | 6 | Specify study characteristics (e.g., PICOS, length of follow-up) and report characteristics (e.g., years considered, language, publication status) used as criteria for eligibility, giving rationale. | 5-6 |
| Information sources | 7 | Describe all information sources (e.g., databases with dates of coverage, contact with study authors to identify additional studies) in the search and date last searched. | 5 |
| Search | 8 | Present full electronic search strategy for at least one database, including any limits used, such that it could be repeated. | Supplementary Table 2 |
| Study selection | 9 | State the process for selecting studies (i.e., screening, eligibility, included in systematic review, and, if applicable, included in the meta-analysis). | 5-6, Fig 1 |
| Data collection process | 10 | Describe method of data extraction from reports (e.g., piloted forms, independently, in duplicate) and any processes for obtaining and confirming data from investigators. | 6 |
| Data items | 11 | List and define all variables for which data were sought (e.g., PICOS, funding sources) and any assumptions and simplifications made. | 6 |
| Risk of bias in individual studies | 12 | Describe methods used for assessing risk of bias of individual studies (including specification of whether this was done at the study or outcome level), and how this information is to be used in any data synthesis. | 6, Supplementary Table 3 |
| Summary measures | 13 | State the principal summary measures (e.g., risk ratio, difference in means). | 6 |
| Synthesis of results | 14 | Describe the methods of handling data and combining results of studies, if done, including measures of consistency (e.g., $I^2$) for each meta-analysis. | 6-7 |
| Risk of bias across studies | 15 | Specify any assessment of risk of bias that may affect the cumulative evidence (e.g., publication bias, selective reporting within studies). | 6, Supplementary Table 3 |
| Additional analyses | 16 | Describe methods of additional analyses (e.g., sensitivity or subgroup analyses, meta-regression), if done, indicating which were pre-specified. | 6-7 |
| **RESULTS** |  |  |  |
| Study selection | 17 | Give numbers of studies screened, assessed for eligibility, and included in the review, with reasons for exclusions at each stage, ideally with a flow diagram. | 8, Fig 1 |
| Study characteristics | 18 | For each study, present characteristics for which data were extracted (e.g., study size, PICOS, follow-up period) and provide the citations. | Table 1 |
| Risk of bias within studies | 19 | Present data on risk of bias of each study and, if available, any outcome level assessment (see item 12). | 8, Table 1 |
| Results of individual studies | 20 | For all outcomes considered (benefits or harms), present, for each study: (a) simple summary data for each intervention group (b) effect estimates and confidence intervals, ideally with a forest plot. | 8-12, Fig 2-12, Table 2-4 |
| Synthesis of results | 21 | Present results of each meta-analysis done, including confidence intervals and measures of consistency. | 8-12, Fig 2-12 |
| Risk of bias across studies | 22 | Present results of any assessment of risk of bias across studies (see Item 15). | 8, 12, Table 1, Fig 11B |
| Additional analysis | 23 | Give results of additional analyses, if done (e.g., sensitivity or subgroup analyses, meta-regression [see Item 16]). | 8-10, 12, Fig 3, 4, 6, 7 |
| **DISCUSSION** |  |  |  |
| Summary of evidence | 24 | Summarize the main findings including the strength of evidence for each main outcome; consider their relevance to key groups (e.g., healthcare providers, users, and policy makers). | 13-17 |
| Limitations | 25 | Discuss limitations at study and outcome level (e.g., risk of bias), and at review-level (e.g., incomplete retrieval of identified research, reporting bias). | 17-19 |
| Conclusions | 26 | Provide a general interpretation of the results in the context of other evidence, and implications for future research. | 19 |
| **FUNDING** |  |  |  |
| Funding | 27 | Describe sources of funding for the systematic review and other support (e.g., supply of data); role of funders for the systematic review. | 1 |

## APPENDIX 2: Search Strategy

| Database Searched | Search Strategy |  |
| --- | --- | --- |
| CINAHL<br><1982 to February 07, 2022> |  |  |
| Ovid Emcare<br><1995 to 2022 Week 4> | 1 | exp head injury/ |
|  | 2 | exp rural health/ or exp rural population/ |
|  | 3 | 1 and 2 |
|  | 4 | limit 3 to (human and English language) |
| Ovid MEDLINE® and Epub Ahead of<br>Print, In-Process, In-Data-Review & Other<br>Non-Indexed Citations, Daily and<br>Versions <1946 to February 04, 2022> | 1 | exp Craniocerebral Trauma/ |
|  | 2 | exp Rural Health/ or exp Rural Population/ |
|  | 3 | 1 and 2 |
|  | 4 | limit 3 to (English language and humans) |
| Scopus<br><1788 to February 07, 2022> | ( "head injur*" OR "head trauma" OR "traumatic brain<br>injur*" OR tbi ) AND ( rural OR remote ) AND ( LIMIT-TO ( DOCTYPE , "ar" ) ) AND ( LIMIT-TO ( SUBJAREA ,<br>"MEDI" ) OR LIMIT-TO ( SUBJAREA , "NEUR" ) ) AND<br>( LIMIT-TO ( LANGUAGE , "English" ) ) AND ( LIMIT-TO ( EXACTKEYWORD , "Human" ) OR LIMIT-TO ( EXACTKEYWORD , "Humans" ) ) |  |

CINAHL, Cumulative Index of Nursing and Allied Health; EBSCO, Elton B. Stephens Company; TBI, traumatic brain injury.

## APPENDIX 3: Modified Newcastle-Ottawa Quality Assessment Scale

| **Selection** |  |
| --- | --- |
| 1 | Is the study population likely to be representative of the whole population? |
| 2 | Was the non-exposed cohort (urban population) sourced from the same state/country or database? |
| 3 | Was the exposure ascertained through secure records, i.e., confirmed medical record? |
| 4 | Does the study specify the source of data? |
| 5 | Was the sample size appropriate for each cohort for statistical comparison of rural and urban populations? |
| 6 | Are inclusion and exclusion criteria clearly outlined? |
| 7 | Is the study population specifically sought for the purpose of the study i.e., not part of a larger shared database? |
| 8 | Is the date range of the data set clearly stated? |
| **Comparability** |  |
| 9 | Is the research methodology clearly stated? |
| 10 | Is the data collection methodology clearly stated? |
| 11 | Is the statistical methodology appropriate? |
| 12 | Does the study report demographics for both rural and urban populations to enable comparison? |
| 13 | Does the study report incidence/prevalence as well as mortality/other outcome measure? |
| 14 | Does the study state the method of determining head injury severity? |
| 15 | Does the study state the method of determining urban and rural classifications? |
| **Outcome** |  |
| 16 | Are the outcomes clearly stated and discussed in relation to the data collection? |
| 17 | Does the study report findings in relation to original aims? |
| 18 | Was follow-up long enough for outcomes to occur without missing data (e.g., mortality, length of stay)? |
| 19 | Were the differences between groups clinically meaningful? (i.e., was the clinical significance reported in addition to statistical significance) |
| 20 | Does the study account for confounding factors and is it considered in the analysis? |

Score \_\_\_\_\_ /20
Low quality <10 Moderate quality 10-15 High quality >15

## APPENDIX 4: MOOSE Checklist for Meta-analyses of Observational Studies

| Item | Recommendation | Reported on Page No |
| --- | --- | --- |
| Reporting of background should include |  |  |
| 1 | Problem definition | 4 |
| 2 | Hypothesis statement | Not applicable |
| 3 | Description of study outcome(s) | 4 |
| 4 | Type of exposure or intervention used | Not applicable |
| 5 | Type of study designs used | Table 1 |
| 6 | Study population | Table 1-2 |
| Reporting of search strategy should include |  |  |
| 7 | Qualifications of searchers (eg, librarians and investigators) | 1 (Title page) |
| 8 | Search strategy, including time period included in the synthesis and key words | 5, Appendix 2 |
| 9 | Effort to include all available studies, including contact with authors | 5 |
| 10 | Databases and registries searched | 5, Appendix 2 |
| 11 | Search software used, name and version, including special features used | 5, Appendix 2 |
| 12 | Use of hand searching (eg, reference lists of obtained articles) | 5 |
| 13 | List of citations located and those excluded, including justification | 8, Fig 1 |
| 14 | Method of addressing articles published in languages other than English | 5 |
| 15 | Method of handling abstracts and unpublished studies | 5 |
| 16 | Description of any contact with authors | 5, 8, Fig 1 |
| Reporting of methods should include |  |  |
| 17 | Description of relevance or appropriateness of studies assembled for assessing the hypothesis to be tested | 5 |
| 18 | Rationale for the selection and coding of data (eg, sound clinical principles or convenience) | 6 |
| 19 | Documentation of how data were classified and coded (eg, multiple raters, blinding and interrater reliability) | 5-6 |
| 20 | Assessment of confounding (eg, comparability of cases and controls in studies where appropriate) | Table 3-4 |
| 21 | Assessment of study quality, including blinding of quality assessors, stratification or regression on possible predictors of study results | 6, Appendix 3 |
| 22 | Assessment of heterogeneity | 6-7 |
| 23 | Description of statistical methods (eg, complete description of fixed or random effects models, justification of whether the chosen models account for predictors of study results, dose-response models, or cumulative meta-analysis) in sufficient detail to be replicated | 6-7 |
| 24 | Provision of appropriate tables and graphics | Table 1-4<br>Fig 1-12<br>Appendices 5-7 |
| Reporting of results should include |  |  |
| 25 | Graphic summarizing individual study estimates and overall estimate | Fig 2-12 |
| 26 | Table giving descriptive information for each study included | Table 1-2 |
| 27 | Results of sensitivity testing (eg, subgroup analysis) | 8-10, 12, Fig 3, 4, 6, 7 |
| 28 | Indication of statistical uncertainty of findings | 8-12 |
| Reporting of discussion should include |  |  |
| 29 | Quantitative assessment of bias (eg, publication bias) | 8, 12, Table 1, Fig 11B |
| 30 | Justification for exclusion (eg, exclusion of non-English language citations) | 5, Fig 1 |
| 31 | Assessment of quality of included studies | 8, Table 1 |
| Reporting of conclusions should include |  |  |
| 32 | Consideration of alternative explanations for observed results | 13-19 |
| 33 | Generalization of the conclusions (ie, appropriate for the data presented and within the domain of the literature review) | 17-19 |
| 34 | Guidelines for future research | 19 |
| 35 | Disclosure of funding source | 1 |

*From:* Stroup DF, Berlin JA, Morton SC, et al, for the Meta-analysis Of Observational Studies in Epidemiology (MOOSE) Group. Meta-analysis of Observational Studies in Epidemiology. A Proposal for Reporting. *JAMA*. 2000;283(15):2008-2012. doi: 10.1001/jama.283.15.2008.

## APPENDIX 5: Data Sources

| Author | Year | Data Source(s) |
| --- | --- | --- |
| Agrawal et al (14) | 2017 | Prospective collection (single tertiary hospital) |
| Andelic et al (16) | 2012 | Analysis of data from prospective population-based multicentre study |
| Asemota et al (26) | 2013 | HCUP-NIS |
| Berry et al (5) | 2010 | AIHW NHMD |
| Brown et al (18) | 2019 | CDC WISQARS™ |
| Chan et al (43) | 2005 | Prospective collection (2 hospitals) |
| Chapital et al (20) | 2007 | Queen's Medical Center trauma database |
| Cheng et al (27) | 2017 | CDC WONDER |
| Cheng et al (45) | 2020 | China DSP |
| Chiang et al (46) | 2006 | Taiwan Head Injury Registry + hospital records (24 hospitals) |
| Chiu et al (19) | 2007 | Prospective collection (26 hospitals) |
| Daugherty et al (28) | 2021 | CDC NVSS |
| Feigin et al (38) | 2013 | CT/MRI records, hospital discharge registers, private hospitals, GP practices, rehabilitation centres, outpatient clinics, coroner/autopsy records, rest homes, community health services, schools, sports centres, ambulance services, prison, ACC database, death certificates, hospital separation data |
| Gabella et al (15) | 1997 | Colorado surveillance system (hospital discharge data + death certificate data) |
| Gontkovsky et al (29) | 2006 | NIDRR TBI Model System + participant/family member interviews |
| Graves et al (30) | 2019 | MarketScan CCAE |
| Halldorsson et al (44) | 2007 | Icelandic hospitals + EDs + healthcare centres + death register |
| Harradine et al (39) | 2004 | Prospective collection (11 rehabilitation units) |
| Harrison et al (40) | 2012 | AIHW NHMD |
| Johnstone et al (31) | 2003 | Missouri Division of Vocational Rehabilitation |
| Karwat et al† (11, 49) | 2009 | Lublin Regional Specialist Hospital records |
| Leonhard et al (32) | 2015 | Oregon Trauma registry |
| Maier et al (17) | 2014 | CT/MRI records (2 hospitals) |
| Ponsford et al (6) | 2012 | Prospective collection (single rehabilitation centre) |
| Pozzato et al (21) | 2019 | NSW Department of Health hospital data |
| Ratliff et al (33) | 2021 | CDC WISQARS™ |
| Reid et al (34) | 2001 | MDH TBI Registry + death certificates |
| Ring et al (41) | 1986 | NSW Hospital Morbidity Statistics + hospital indices (133 hospitals) + death certificates |
| Robertson & McConnel (35) | 2011 | Children's Medical Center Dallas trauma census |
| Schootman & Fuortes (36) | 2000 | NAMCS & NHAMCS |
| Simpson et al (42) | 2016 | Prospective collection (11 rehabilitation units) + NSW electronic database |
| Stewart et al (37) | 2014 | NACRS |
| Tesfaw et al (47) | 2021 | Prospective collection (single tertiary hospital) |
| Woodward et al (22) | 1984 | South Australian Health Commission hospital separation discharges/transfers/deaths |
| Yates et al (48) | 2006 | Royal Devon and Exeter Hospital ED database |

† Karwat et al (2009A and 2009B) report different outcomes on the same patient population, and are therefore considered as one study. HCUP-NIS, Healthcare Cost and Utilization Project Nationwide Inpatient Sample; AIHW NHMD, Australian Institute of Health and Welfare National Hospital Morbidity Database; CDC, Centers for Disease Control and Prevention; WISQARS™, Web-based Injury Statistics Query and Reporting; WONDER, Wide-ranging Online Data for Epidemiological Research; DSP, Disease Surveillance Points; NVSS, National Vital Statistics System; CT, computed tomography; MRI, magnetic resonance imaging; GP, General Practitioner; ACC, Accident Compensation Corporation; NIDRR, National Institute in Disability and Rehabilitation Research; TBI, traumatic brain injury; CCAE, Commercial Claims and Encounters database; ED, emergency department; NSW, New South Wales; MDH, Minnesota Department of Health; NAMCS, National Ambulatory Care Survey; NHAMCS, National Hospital Ambulatory Care Survey; NACRS, National Ambulatory Care Reporting System.

## APPENDIX 6: Inclusion and Exclusion Criteria

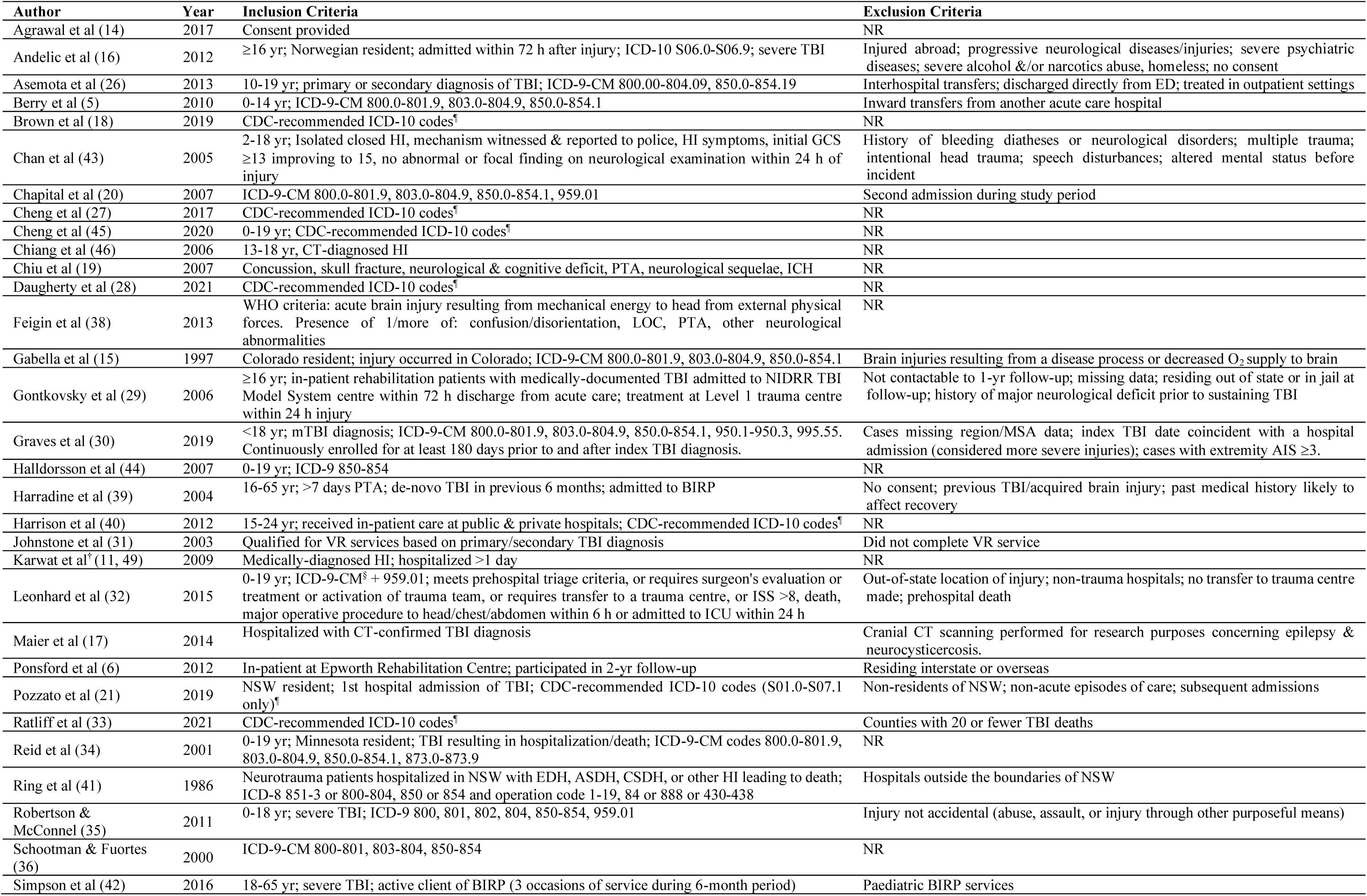
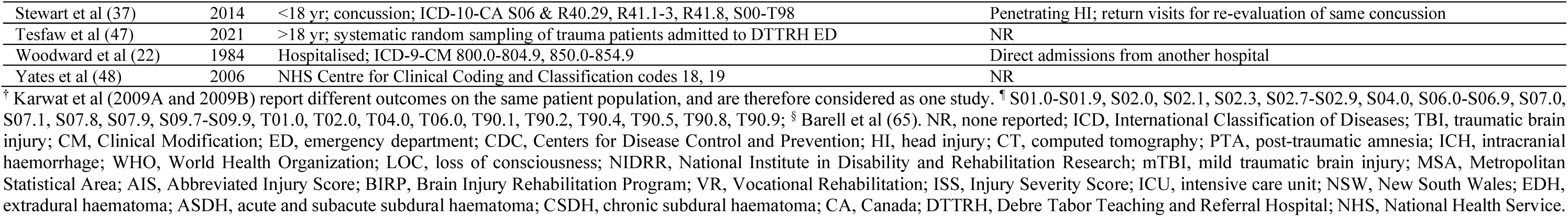

## APPENDIX 7: Rural/Urban Classifications

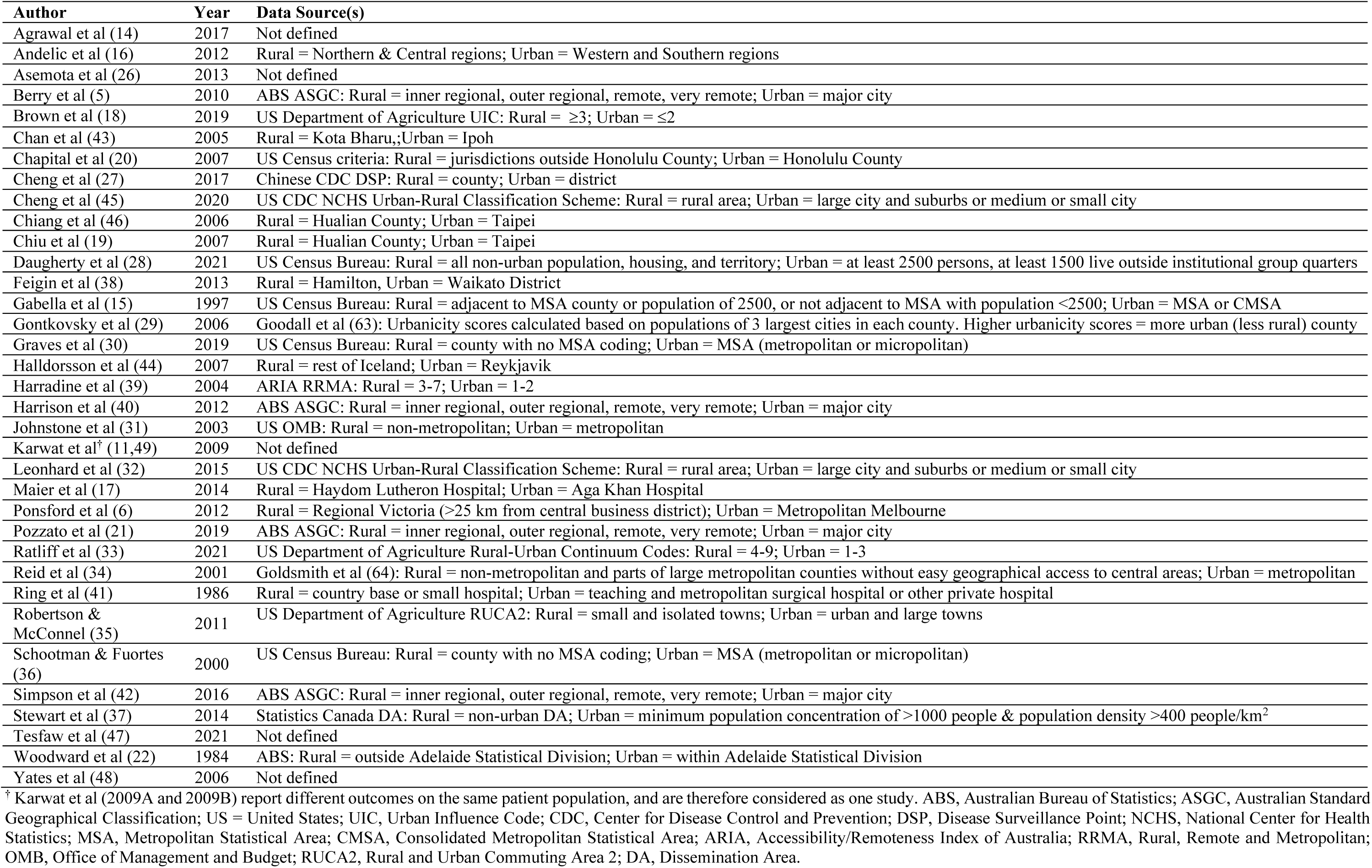

